# The Gene Expression Landscape of Disease Genes

**DOI:** 10.1101/2024.06.20.24309121

**Authors:** Judit García-González, Alanna C. Cote, Saul Garcia-Gonzalez, Lathan Liou, Paul F. O’Reilly

## Abstract

Fine-mapping and gene-prioritisation techniques applied to the latest Genome-Wide Association Study (GWAS) results have prioritised hundreds of genes as causally associated with disease. Here we leverage these recently compiled lists of high-confidence causal genes to interrogate where in the body disease genes operate, which, in previous studies, has mostly been investigated by testing for enrichment of GWAS signal among genes with cell/tissue specific expression. By integrating GWAS summary statistics, gene prioritisation results, and RNA-seq data from 46 tissues and 204 cell types, we directly analyse the gene expression of putative disease genes across the body in relation to 11 major diseases and cancers. In tissues and cell types with established disease relevance, disease genes show higher and more specific gene expression compared to control genes. However, we also detect elevated expression in tissues and cell types without previous links to the corresponding disease. While some of these results may be explained by cell types that span multiple tissues, such as macrophages in brain, blood, lung and spleen in relation to Alzheimer’s disease (*P*-values < 10^-3^), the cause for others is unclear and warrants further investigation. To support functional follow-up studies of disease genes, we identify technical and biological factors influencing their expression, and highlight tissues in which higher expression is associated with increased odds of inclusion in drug development programs. We provide our systematic testing framework as an open-source, publicly available tool that can be utilised to offer novel insights into the genes, tissues and cell types involved in any disease, with the potential for informing drug development and delivery strategies.

## Introduction

Genome-wide association studies (GWAS) have identified thousands of risk loci for complex diseases over the past two decades^1^. An important aspect of translating these findings into biological and clinical insights is pin-pointing the causal variants and genes. To this end, fine-mapping and gene prioritisation strategies have been developed ^2^ and − only in recent years − lists of high-confidence causal genes have been compiled across multiple diseases ^3–11^. These curated lists of putative disease genes provide the opportunity to systematically and directly examine where in the body disease genes are active, offering insights into disease initiation, aetiological mechanisms and therapeutic opportunities. For example, successful drug targets are enriched for GWAS prioritised genes^12^, as well as tissue specific genes^13–15^. This suggests that mapping the tissues and cell types in which disease genes are active could further enhance the identification of new drug targets ^16,17^ with reduced off-target effects^18,19^.

Previous studies integrating GWAS and gene expression data to infer the tissues/cells involved in disease have done this more indirectly. Typically, such studies identify genes with the greatest relative (i.e. specific) expression in each cell type or tissue, irrespective of disease, and then assess their enrichment in GWAS signals for the disease of interest ^20–23^. While this approach provides insight into the tissues and cell types where disease may initiate or progress, it does not directly capture the question of most interest: *how and where disease genes are expressed*. Existing methods do not leverage lists of high-confidence causal disease genes, and, thus, they include many genes with no involvement in the disease under study in their testing and cannot explore how different definitions of disease genes influence the identification of disease-relevant tissues and cell types. Moreover, most current approaches focus on relative gene expression (i.e. specificity) only. Although investigating relative expression may highlight disease genes less likely to have pleiotropic effects, absolute gene expression, capturing overall abundance, may be more important for diseases that involve multiple cell/tissue types. Thus, while a gene expressed at high levels across multiple tissues could clearly play a pivotal role in a disease, it would be missed by methods restricted to tissue-specific signal.

In this study, we use three alternative approaches that leverage GWAS and RNA-seq data to infer where disease genes are expressed: the first − that we call “*GWAS to Gene Expression”* − is a novel approach that tests whether putative disease genes, according to recent large-scale GWAS, exhibit distinct expression levels across the body compared to control genes (**Fig 1a**). We ensure the robustness of this approach by employing three distinct definitions for disease genes and utilising three different sets of control genes. The second approach − that we call “*Gene Expression to GWAS”* − examines whether high-expression genes are enriched for GWAS signal, as calculated by MAGMA^22^ (**Fig 1b**). In the third approach, we perform a systematic PubMed search to assess the evidence for tissue-disease associations reported in the literature (**Fig 1c**). Contrasting results systematically across the three approaches allows for a degree of ‘triangulation’ of results. We apply our testing framework in 46 tissues and 204 cell types obtained from the GTEx^24^, ARCHS4^25^ and Tabula Sapiens^26^ resources (**Fig 1d**). We also characterise, for each individual gene, to what extent different predictors (batch ID, subject ID, age, sex etc) contribute to gene expression (**Fig 1e**) and examine how both absolute and tissue-specific (relative) expression of disease genes influence their odds for being included in drug development programs (**Fig 1f**). Analyses are performed across eight major diseases and three cancers, for which powerful GWAS and curated lists of high-confidence disease genes are available: Schizophrenia (SCZ), Inflammatory Bowel Disease (IBD), Alzheimer’s Disease (AD), Coronary Artery Disease (CAD), Bipolar Disorder (BD), Type 2 Diabetes (T2D), Attention-Deficit/Hyperactivity Disorder (ADHD), Serum 25 Hydroxyvitamin D (Vitamin D), and Breast, Prostate and Colorectal cancer.

**Figure 1.**
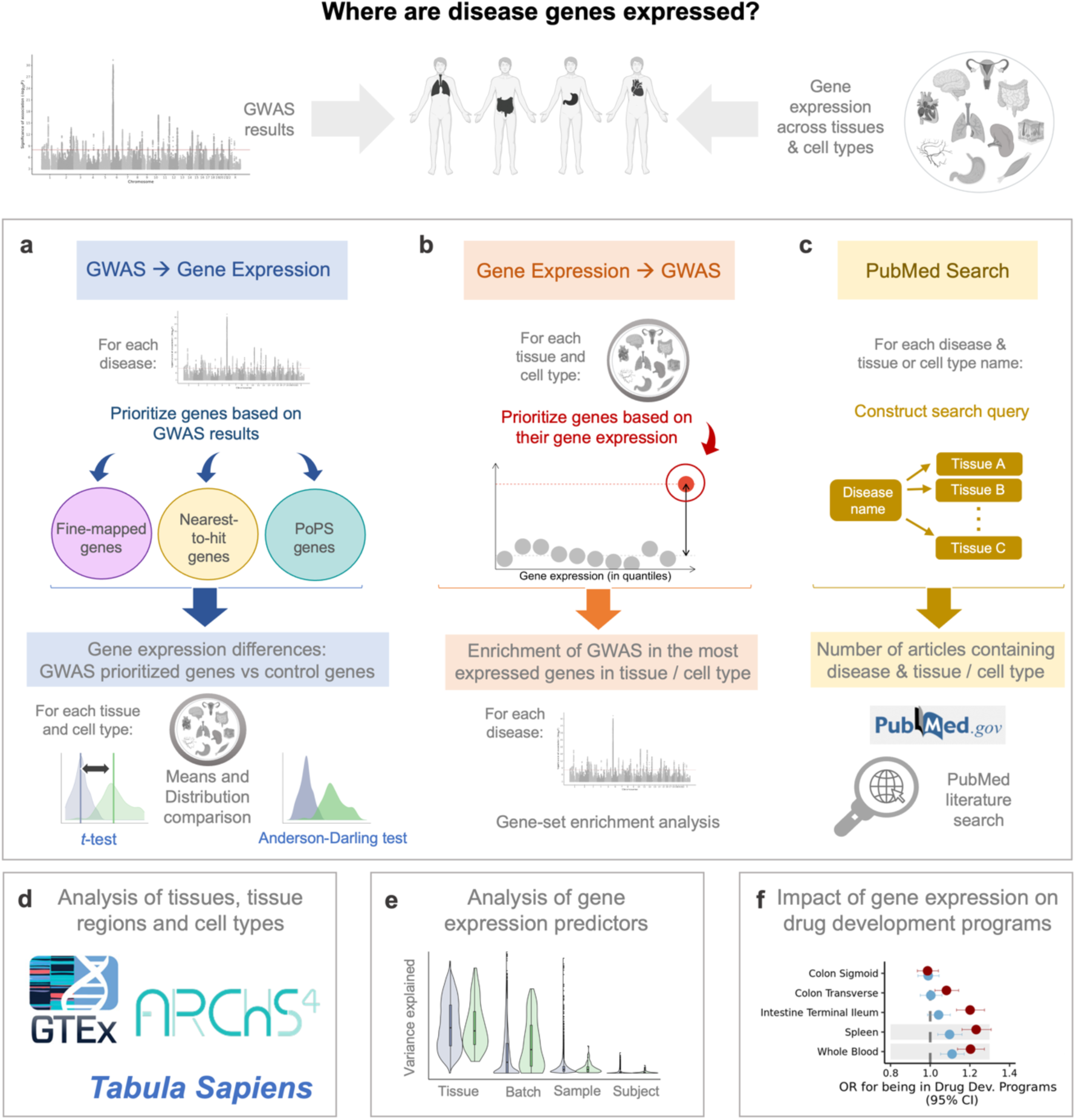
Study overview. **a**, Approach using lists of prioritised, high-confidence disease genes, derived from GWAS results. **b**, Approach using genes with the highest absolute or relative expression in each cell and tissue. **c**, PubMed-based literature search to assess the tissues most often cited in relation to each disease. **d**, Primary analyses apply approaches a-c to the tissue gene expression data of GTEx, while follow-up analyses are applied to the cell types of the ARCHS4 and Tabula Sapiens datasets. **e**, Factors affecting gene expression variance are analysed for each disease gene. **f**, For tissues where disease genes are significantly more expressed than control genes (*t*-test, Bonferroni-corrected), we assess how absolute and relative expression of disease genes influence their likelihood of inclusion in drug development programs. Figure partially created using BioRender.com.

## Results

### Exploring the expression profiles of disease genes

To infer where in the body disease genes operate, here we leverage GWAS and gene expression data utilising three alternative strategies of inference: (i) *‘GWAS to Gene Expression’*; (ii) *‘Gene Expression to GWAS’*; and (iii) *‘PubMed Search’*. The first part of the results presents our primary analyses, which examine gene expression in tissues across the three strategies. We then extend these analyses to the cell-type level to refine our findings within relevant tissues.

#### GWAS to Gene Expression

A range of statistical and computational approaches have been developed^27–31^ to link GWAS-associated variants to their target genes and to prioritise genes based on GWAS results. Given the absence of a single gold standard approach for gene prioritisation, we employ and describe below three alternative approaches to define ‘disease genes’ inferred from GWAS results: (i) *nearest-to-hit genes*, (ii) *fine-mapped genes*, and (iii) *PoPS genes*. All remaining protein-coding genes are used as control genes (**Methods**).

*(i) Nearest-to-hit genes*: While sophisticated methods to prioritise causal genes exist (see below), previous research suggests that the nearest gene to the lead variant is far more likely to be the causal gene than others at the locus ^27,31,32^. We performed clumping on the GWAS hits and identified an average of 162 ‘nearest-to-hit’ genes for each disease/trait, with an average distance of 28 kb between the SNP and the closest protein coding gene (See **Methods, Table 1** and **Supplemental Table 1**).
*(ii) Fine-mapped genes*: The latest landmark GWAS of the major diseases^3–11^ have incorporated sub-studies performing a range of gene prioritisation analyses to produce lists of around 50-300 high-confidence causal genes for each disease. While different studies use a different selection of methods to prioritise putatively causal genes, most of them integrate GWAS results with the latest functional genomics data (**Methods**). Our literature search on fine-mapped genes resulted in gene lists ranging from 49 genes prioritised for BD to 281 genes prioritised for IBD, with an average of 126 genes across the 8 diseases investigated (**Supplemental Table 2**).
*(iii) PoPS genes:* The method ‘Polygenic Priority Scores’ (PoPS) method^31^ leverages gene-level *Z*-scores from GWAS (calculated using the software MAGMA^22^), as well as gene features from single-cell gene expression data, biological pathways and predicted protein-protein interaction networks to prioritise putatively causal disease genes. For each trait, we extracted the genes with the top 1% PoPS scores, corresponding to 184 protein-coding genes with the highest PoPS scores for each trait. The top 1% threshold was chosen because it provides a similar number of genes as the other two prioritisation approaches, limiting biases due to differences in the number of genes included for each group of putative disease genes (**Supplemental Table 3**).

**Table 1.** Gene prioritisation based on the nearest gene to GWAS hit. Table shows references for each GWAS summary statistics, the number of clumped SNPs, genes and median distance between SNP and selected gene. CAD, Coronary Artery Disease; SCZ, Schizophrenia; IBD, Inflammatory Bowel Disease; AD, Alzheimer’s Disease; BD, Bipolar Disorder; ADHD, Attention-deficit/hyperactivity disorder; T2D, Type 2 Diabetes.

| Trait | GWAS summary statistics used | N clumped SNPs | N unique ensemble IDs | Median distance between SNP and nearest protein coding gene (bp) |
| --- | --- | --- | --- | --- |
| CAD | Aragam et al, 2022<sup>3</sup> | 525 | 297 | 14,734 |
| SCZ | Trubetskoy et al. 2022<sup>4</sup> | 452 | 310 | 18,125 |
| IBD | Liu et al, 2015<sup>50</sup> | 487 | 285 | 11,724 |
| AD | Bellenguez et al, 2022<sup>11</sup> | 239 | 99 | 8,363 |
| BD | Mullins et al, 2021<sup>51</sup> | 73 | 63 | 20,660 |
| ADHD | Demontis et al, 2023<sup>8</sup> | 32 | 22 | 104,008 |
| T2D | Suzuki et al, 2023<sup>52</sup> | 50 | 31 | 26,633 |
| Vitamin D | Revez et al, 2020<sup>6</sup> | 517 | 200 | 22,111 |

For each disease, we performed *t-*tests comparing the absolute and relative expression of high-confidence disease genes (fine-mapped, nearest-to-hit and PoPS genes) with those of the control genes. Disease genes show higher expression in tissues with known relevance to the corresponding disease (**Fig 2,** *t-*test *P*-values shown in blue columns). Overall, the disease genes’ relative gene expression *t-*test results are similar to those for absolute gene expression but generally have smaller *P*-values. Unless otherwise noted, the results and *P*-values discussed below refer to absolute gene expression.

**Figure 2.**
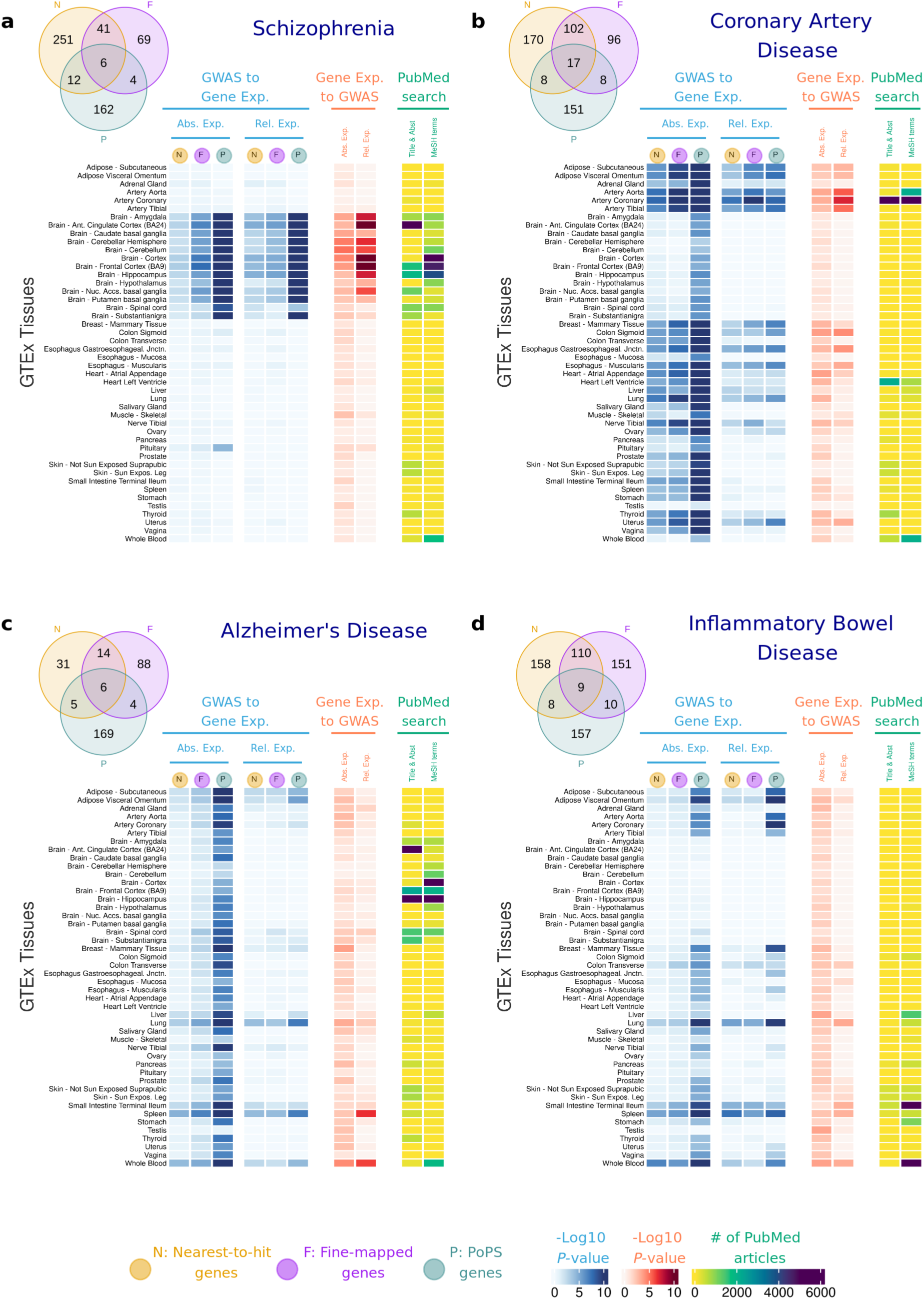
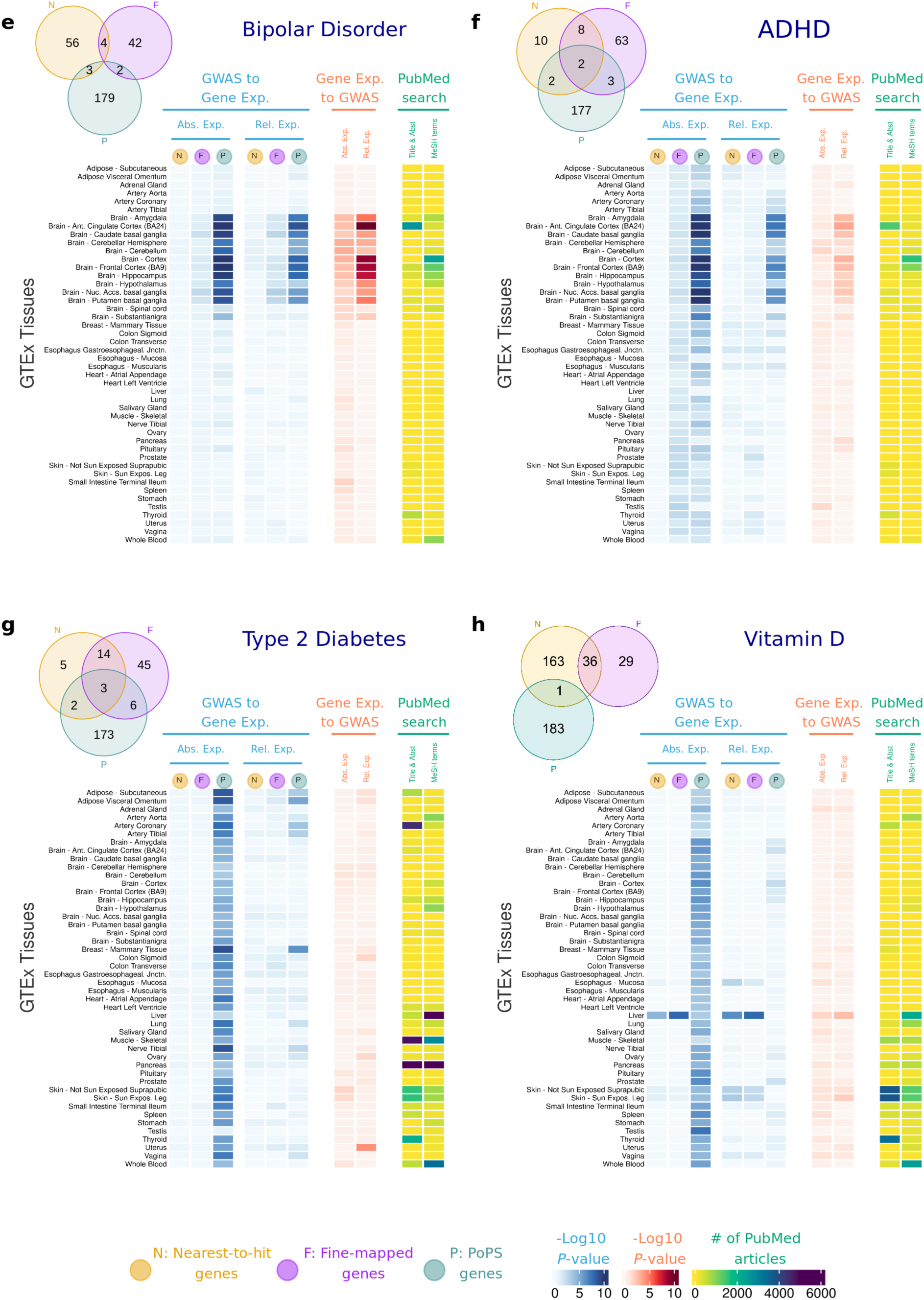
Heatmap showing results of the association between gene expression in each GTEx tissue and **a**, Schizophrenia; **b**, Coronary Artery Disease; **c**, Alzheimer’s Disease; **d**, Inflammatory Bowel Disease; **e**, Bipolar Disorder; **f**, ADHD; **g**, Type 2 Diabetes; **h**, Vitamin D. In blue, results showing the Log10 *P-*value for a one-side *t-*tests, testing the null hypothesis that disease-associated genes are not more expressed than other protein-coding genes expressed in that tissue. In red, results showing the Log10 *P-*value for enrichment of GWAS signal across the set of genes with highest absolute and relative expression for each tissue. In yellow, results for the Literature Search using PubMed. Abs. Exp, Absolute expression; Rel. Relative expression; F, Fine-mapped genes; N, Nearest-to-hit genes, P, Polygenic Priority Scores genes; Title & Abst, Title and Abstract; MeSH, MeSH terms.

SCZ genes are more expressed in the brain, although *P-*values vary substantially across brain tissues (from *P-*value = 3.1×10^-2^ for the nearest gene list in substantia nigra to *P-*value = 2.0×10^-29^ for PoPS genes in the brain cortex). CAD genes are more expressed in the aorta, coronary and tibial arteries (*P-*values < 10^-7^). IBD genes are most expressed in the small intestine and colon transverse (*P-*values < 10^-4^ except for nearest-to-hit genes). Vitamin D genes are most expressed in the skin and in the liver, with the latter tissue being where the biologically inactive vitamin D_3_ is activated to produce 25-hydroxyvitamin D_3_ ^33^.

Other significant results point to tissues not typically linked to the disease: AD genes show highest expression in the blood, spleen (PoPS genes *P-*value < 10^-13^, nearest-to-hit genes *P-* value < 10^-5^) and adipose tissues (PoPS genes *P-*values < 10^-13^) but not in the brain. SCZ genes are highly expressed in the pituitary (PoPS genes *P-*values = 3.7×10^-5^). CAD genes exhibit higher expression across multiple tissues including reproductive, adipose, and digestive systems, as well as in the lung (*P-*values < 10^-6^). IBD genes have highest expression in lung (*P-* values < 10^-4^), blood and spleen (*P-*values < 10^-6^). Differences between T2D genes vs control genes appear most significant in breast mammary tissues (*P-*values < 10^-7^), but these results are not driven by differences between males and females, or the presence of some genes with high expression in only one sex (**Additional File 1, Supplemental Material** and **Supplemental Figure 1**).

We also applied the Anderson-Darling test^34^ to evaluate whether the expression *distribution* of high-confidence disease genes differs from that of control genes. In most cases, results were consistent with the *t*-tests but yielded smaller *P-*values (**Supplemental Figure 2**). Full results for both tests, across all diseases, tissues and gene lists are provided in **Supplemental Table 4**. We performed several sensitivity analyses to ensure the robustness of our findings. First, we tested the impact of varying the PoPS top percentage threshold (**Methods** and **Supplementary** Figure 3). Second, we examined alternative definitions of control groups, repeating our analyses using genes prioritised by the Open Targets initiative^41^ (**Methods**, **Supplemental Figures 4-11** and **Supplemental Table 5**). Third, we investigated whether the observed results could be explained by a small number of genes with extremely high expression. We repeated the analyses of Fig.2 excluding genes in the top 10% of absolute and relative expression for tissues with significant results. (**Additional File 1, Supplemental Material, Supplemental Table 6**). Results show that, while the *P-*values increase for all the tests, results are qualitatively consistent (**Supplemental Figures 12-13** and **Supplemental Table 7**). For example, the cortex and cingulate cortex are significantly associated with SCZ (Absolute expression *P*-values < 10^-20^), and the tissues most associated with AD remain the small intestine (*P*-value = 3.8×10^-9^), spleen (*P*-value = 4.9×10^-8^) and lung (*P*-value = 5.0×10^-8^).

#### Gene Expression to GWAS

In this section, we test whether genes with high absolute and relative expression in a tissue are enriched for GWAS signal. In an early example of this approach, Skene et al.^35^ assessed the enrichment of GWAS signal using MAGMA^23^ and LDSC ^21^, among genes in the top decile of absolute and relative (also called specific) gene expression (see **Methods**). We expand the approach here to investigate *absolute gene expression*, in addition to *relative gene expression*. Our findings are overall consistent with the results of the GWAS to Gene Expression approach. (**Figure 2**, red columns). For SCZ, BP and ADHD, MAGMA results were nominally significant in numerous brain tissues (*P*-value < 0.05), such as cortex, anterior cingula, hippocampus, amygdala, or cerebellum or nucleus accumbens. The strongest results were observed for the cortex and anterior cingula tissues and SCZ (*P*-value < 10^-12^). For CAD, numerous tissues show significant *P*-values for both absolute and relative expression, with arteries (*P*-value < 10^-6^), colon sigmoid (*P*-value = 4.7×10^-6^) and esophagus (*P*-value = 8.4×10^-5^) having the strongest enrichment of GWAS signal among their most specific genes. In IBD, the intestine, blood, testis, liver, lung, and spleen are the tissues with strongest enrichment (*P*-values < 10^-3^), while AD shows the strongest result in spleen and blood (*P*-values < 10^-6^). For Vitamin D, liver is the most relevant tissue (*P*-value = 2.5×10^-3^). For T2D, none of the tissues showed significant results. Overall, MAGMA enrichments for relative gene expression are more pronounced than for absolute gene expression. MAGMA results for each disease, each tissue, and each gene list are included in **Supplemental Table 8**.

#### PubMed Search

Next, we investigated disease-tissue associations by performing a systematic literature search via PubMed, using two approaches to construct PubMed search queries. First, we use Medical Subject Headings (MeSH) terms, a standardized vocabulary from the National Library of Medicine. Second, we identify tissue/disease pair names in the title and abstract of the PubMed articles. These two approaches provide consistent results (**Figure 2**, yellow columns). While the PubMed search results largely support our findings, there are tissues identified in our main analyses that may be understudied, as the support in the literature is low (e.g. spleen in AD and IBD, digestive tissues in CAD). There are also disease-tissue pairs that have been well-studied (e.g. Skeletal muscle and Vitamin D, or Liver and Inflammatory Bowel Disease), but that have limited support in our analyses. Results with the number of papers found for each query are included in **Supplemental Table 9 & 10**.

#### Correlation in results across the three alternative strategies

Across the different sets of tests, we observe strong correlations between the *t-*tests and Anderson-Darling tests (mean r = 0.73) and between *t-*tests and MAGMA (mean r = 0.55), with most P-values < 1×10^-3^. Correlations across the three approaches varied by disease, with the highest correlations in SCZ (r = 0.83), CAD (r = 0.71), and IBD (r = 0.69), while T2D exhibited the lowest correlation (r = 0.27). Detailed results are provided in **Supplemental Table 11**.

### The gene expression landscape at the cell-type level

To further investigate where in the body disease genes operate, we repeated our testing framework in a set of cell types and tissue regions extracted from the (i) Tabula Sapiens^26^, a resource combining data on nearly 500,000 cells from 24 different tissues and organs, many from the same donor, and (ii) ARCHS4^25^, a resource that aggregates RNA-seq data from the Gene Expression Omnibus and the Sequence Read Archive. Gene expression in cell types may have affected the results of our tissue-level analyses because the same cell types are found across different tissues and tissue-level expression is more likely to reflect abundant, rather than rare, cell types.

Figure 3 presents results of the analyses conducted across tissues from Tabula Sapiens for Vitamin D, IBD and CAD, and Figure 4 presents results of the analyses for AD using tissue regions and cell-type data from ARCHS4 (Figure 4a) and Tabula Sapiens (Figure 4b). Results for the other diseases using ARCHS4 can be found in **Supplemental Figure 13** and **Supplemental Tables 12-14**. We focus on Vitamin D, AD and IBD here because significant results were observed in unexpected tissues, such as the spleen and lung, and on CAD because it shows the largest number of associated tissues.

**Figure 3.**
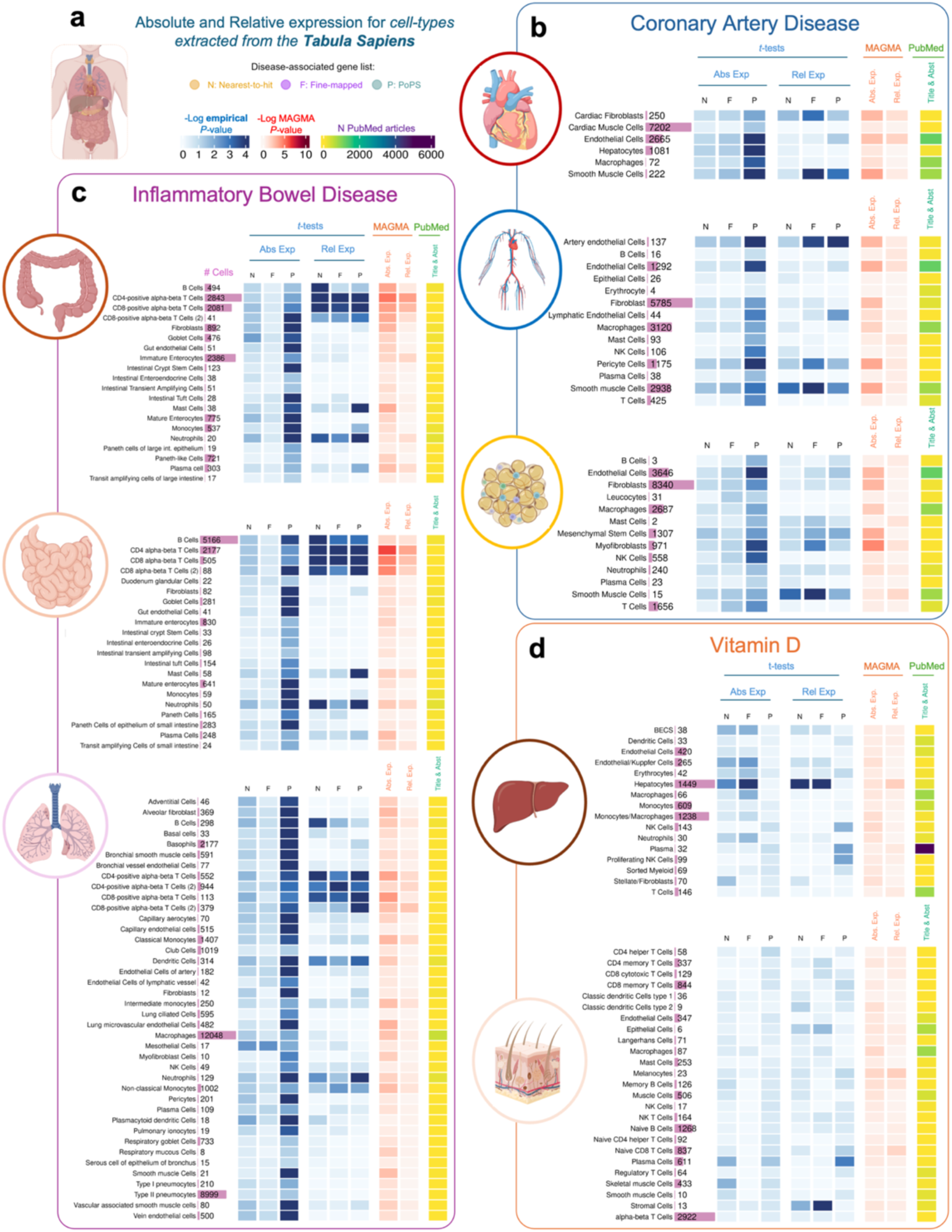
Heatmap showing results of the association between gene expression in each cell type obtained from the Tabula Sapiens dataset **a**, Tabula Sapiens datasets were downloaded for each tissue, and absolute and relative expression was calculated for each cell type in each tissue. **b**, Results for Coronary Artery Disease. **c**, Results for Inflammatory Bowel Disease. **d**. Results for Vitamin D. In blue, results showing the Log10 empirical *P-*value after running 10,000 permutations, testing the null hypothesis that disease-associated genes are not more expressed than other protein-coding genes expressed in that tissue. In red, results showing the Log10 *P-*value for enrichment of GWAS signal across the set of genes with highest expression for each tissue. In yellow, results for the Literature Search using PubMed. Abs. Exp, Absolute expression; Rel. Exp, relative expression; F, Fine-mapped genes; N, Nearest-to-hit genes, P, Polygenic Priority Scores genes; Title & Abst, Title and Abstract are used in the PubMed Search.

**Figure 4.**
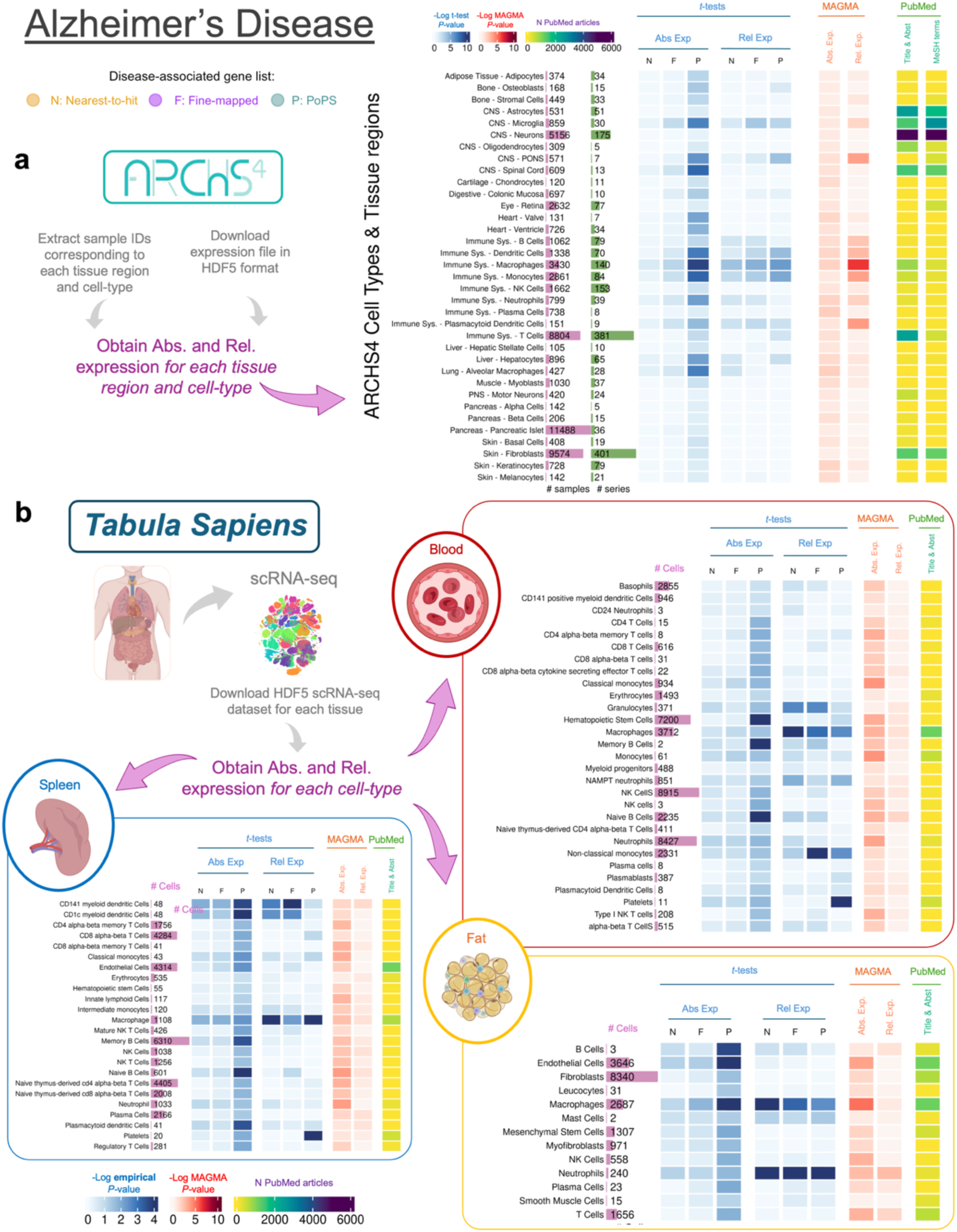
Heatmap showing results of the association between gene expression in each cell type obtained from the Tabula Sapiens dataset. **a**, Results using RNA-seq data from cell types and tissue regions extracted from the ARCHS4 resources. **b**, Results using RNA-seq data from the Tabula Sapiens for blood (red section), spleen (blue section), and fat (yellow section).

Overall, the results for ARCHS4, Tabula Sapiens and GTEx are consistent, although the *P*-values for all cell-type analyses tend to be larger. In the Tabula Sapiens, PoPS genes typically show higher expression than control genes, but these results are often not replicated in fine-mapped genes or nearest-gene lists (**Supplemental Tables 15-17**). Therefore, only results that replicate in at least two gene lists are discussed below.

*GWAS to Gene Expression* results for IBD in cell types derived from the small intestine, large intestine, and lung show that IBD genes present higher relative expression in T-cells (*P*-values for relative expression < 0.02), B cells (*P*-values < 0.038), neutrophils (*P*-values < 0.034) and dendritic cells (*P*-values < 0.01). Results from the *gene expression to GWAS* analyses were largely consistent with these findings.

For CAD, the most significant differences between disease and control genes were observed for relative expression of T-cells in the vasculature system (*P*-value = 0.0028), smooth muscle cells (*P*-value < 0.027) and cardiac fibroblasts (*P*-value < 0.045) in the heart, and fat tissues (*P*-value < 0.044 for fine-mapped genes, *P*-value < 0.014 for PoPS and nearest genes). Various immune cell types, such as macrophages, mast cells, and NK cells in the vasculature system were also significant (*P*-value < 0.015) but only for PoPS genes.

For Vitamin D, the results aligned with the GTEx and ARCHS4 analyses. We observed the most significant difference in expression in the liver, specifically in hepatocytes, between disease-associated and control genes (P-values < 0.0012).

Figure 4a shows results for AD analyses using ARCHS4, showing high absolute and relative expression of AD genes in immune cell types, including dendritic cells, macrophagues and neutrophils (e.g. *P*-values < 0.05). Figure 4b shows similar findings using Tabula Sapiens, with significant expression in macrophages (in the spleen, blood, and fat) and neutrophils (in fat). Due to the lack of brain tissue data in Tabula Sapiens, brain-related cell types were examined only in the ARCHS4 dataset. Microglia − known to play a critical role in AD^36^ − is the only brain tissue significantly associated for both absolute (*P*-value < 0.005) and relative expression (*P*-value < 0.05).

For SCZ, BP, and ADHD, disease genes show significant absolute and relative expression in motor neurons in ARCHS4 (*P*-value = 0.00021 for PoPS genes; *P*-value = 0.036 for fine-mapped genes), but not in the broader category of neurons, despite their larger sample size (**Supplemental Figure 13**).

#### The gene expression landscape of cancer genes

We extended our analysis framework to test the gene expression landscape of eight common cancer types (**Additional File 1, Supplemental Materials**). For three of the cancers, breast, prostate, and colorectal cancer, we applied our full testing pipeline across GTEx, ARCHS4 and Tabula Sapiens datasets since robust GWAS, prioritised gene lists, and high PoPS score genes were available. For five additional cancers (bladder, kidney, lung, ovarian, and pancreatic cancer), no gene prioritisation analyses had been performed^37^, so we tested the “*GWAS to gene expression*” approach using the ‘nearest-to-hit’ method on GTEx. Since cancers have well-established primary tissues of origin (e.g., the colon for colorectal cancer), these analyses serve as a positive control for benchmarking the different approaches and tests.

For the three cancer types with full prioritised gene lists, GTEx results show the most significant and consistent associations with their respective cancer-relevant tissues (P-values < 10^-8^): mammary tissue for breast cancer (**Supplemental Figure 14)**, prostate for prostate cancer (**Supplemental Figure 15**), and colon transverse for colorectal cancer (**Supplemental Figure 16**). Additional significant associations are observed in other tissues, with cell-type level analyses from ARCHS4 and Tabula Sapiens providing further resolution. For example, in prostate cancer, epithelial cells from prostate tissue (luminal, CPE-high luminal, and CGB3A1-high club) had the strongest associations (*P*-values < 10^-4^). For breast cancer, results vary by gene prioritisation method, but B cells and mature luminal cells in the Tabula Sapiens mammary tissue were consistently implicated in both the “*GWAS to Gene Expression*” and the “*Gene Expression to GWAS*” approaches. In colorectal cancer, fibroblasts, monocytes and neutrophils show the most significant associations, supporting the role of immune cells in the disease mechanism. Results from analyses stratified by sex are similar (**Supplemental Figure 17**). No significant associations are observed for the five cancers in which we employed the ‘nearest-to-hit’ prioritisation method, probably due to the low number of genes available for each analysis: 12 genes were prioritised for bladder, 13 for kidney, 16 for lung, 22 for pancreas and 28 for ovary (**Supplemental Figure 18**).

### Predictors of gene expression of disease genes

To characterise the biological factors (such as tissue, sample ID, subject ID) and technical factors (such as batch ID) that contribute to variability in the gene expression of each high-confidence disease gene, we used the ’*variancePartition*’ R package (v.4.3)^38^ (**Methods**).

Disease genes show considerable variability in the main factors that contribute to expression variability (Figure 5a and **Supplemental Table 18**): for some, tissue type explains more than 70% of the expression variance, whereas for others, factors such as batch or collection method are the primary predictors, and tissue type accounts for less than 20% of the variance. Figure 5b**, c** shows schizophrenia as an example, and **Supplemental Figures 19-21** and the R Shiny website associated with this manuscript https://juditgg.shinyapps.io/diseasegenes/ include gene-level results quantifying the contribution of each variable to the variation in expression of each gene and disease.

**Figure 5.**
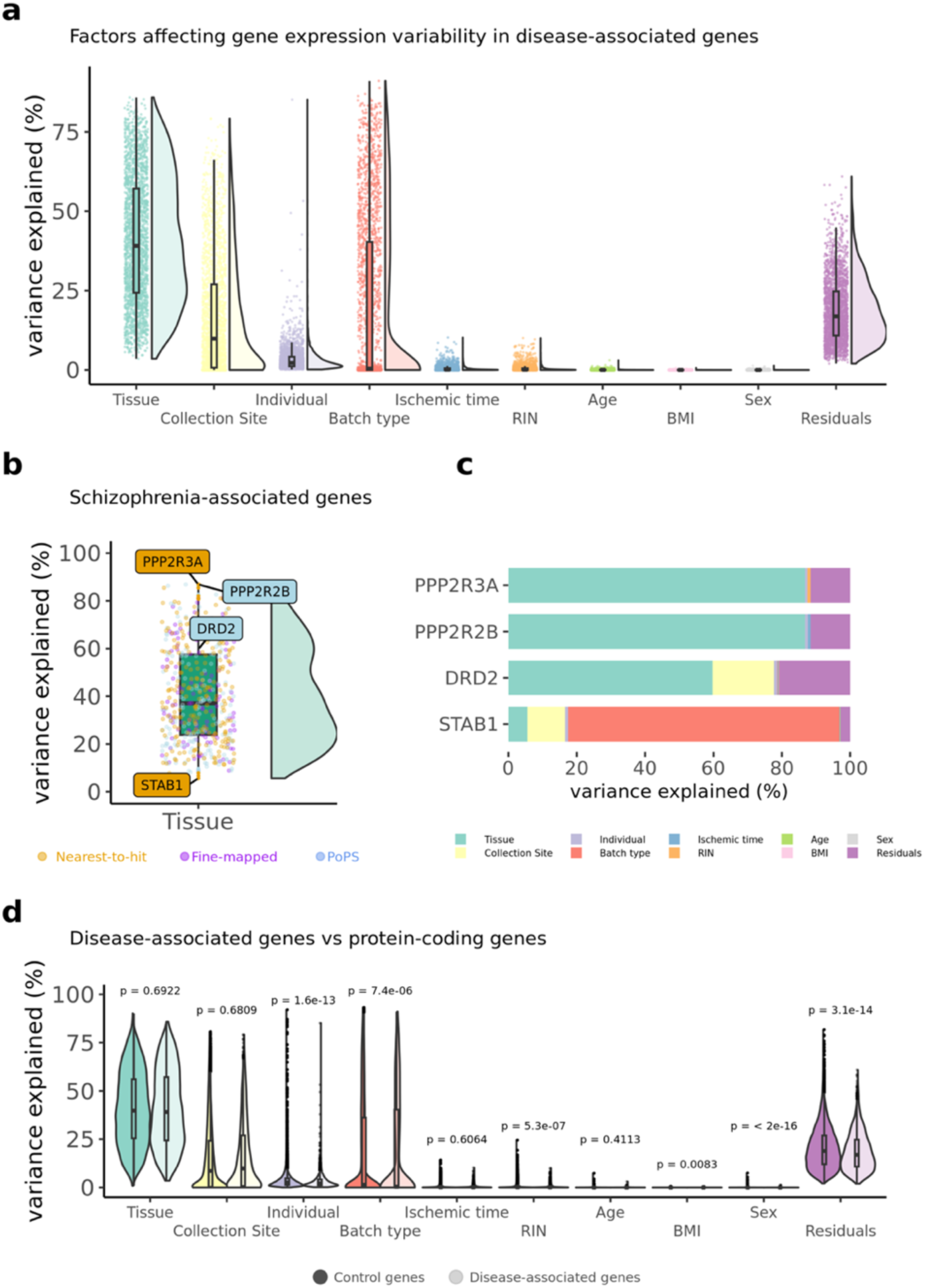
Variance partition is used to investigate the factors influencing gene expression variability in disease-associated genes. **a**, Violin plots representing the distribution of variance partition across all disease-associated genes for the eight diseases investigated. **b**, variance partition results for genes associated with schizophrenia. Genes labelled represent: two genes encoding protein phosphatases (PPP2R3A & PPP2R2B) where tissue-type explain a large fraction in gene expression variance, and a gene (STAB1) where tissue explains less than 10% in gene expression variance. The dopamine receptor 2 (DRD2) is also included because it is the main receptor for most antipsychotic drugs^39,40^. **c**, Bar plots of individual genes showing the variance partition estimates at the individual gene level for genes highlighted in panel b, **d**, Violin plots showing the differences in variance partition results between disease-associated genes and control genes.

Given the gene-level differences observed, we tested whether *variancePartition* results differ between high-confidence disease genes and control genes (Figure 5d). Results were consistent across individual gene lists and diseases, except for Vitamin D, where tissue type explained more variance in disease genes than in control genes (**Supplemental Figs 22-24**).

### Impact of gene expression on the inclusion of disease genes in drug development programs

Previous research has demonstrated that genes with support from GWAS are more likely to become successful drug targets ^12,16^, important given the high failure rates and associated substantial financial losses of the drug discovery process^41^. Similarly, genes with higher cell and tissue-type expression have been found to be more likely to enter drug development programs^42^ and succeed as drug targets^19^. However, no studies to date have examined the association between gene expression and drug development success among disease genes prioritised by GWAS.

For each tissue, we conducted logistic regression analyses using gene expression levels as independent variables and the presence (yes/no) of each gene in drug development programs as the dependent variable. These regressions were performed for tissues in which disease genes exhibited significantly higher expression than control genes after Bonferroni correction. (referred as ‘*GTEx Tissues relevant to the disease*’ in Figure 6). Gene–drug target pairs and their corresponding presence in drug development programs were extracted from a dataset with all target–indication pairs since 2000 (**Methods**).

**Figure 6.**
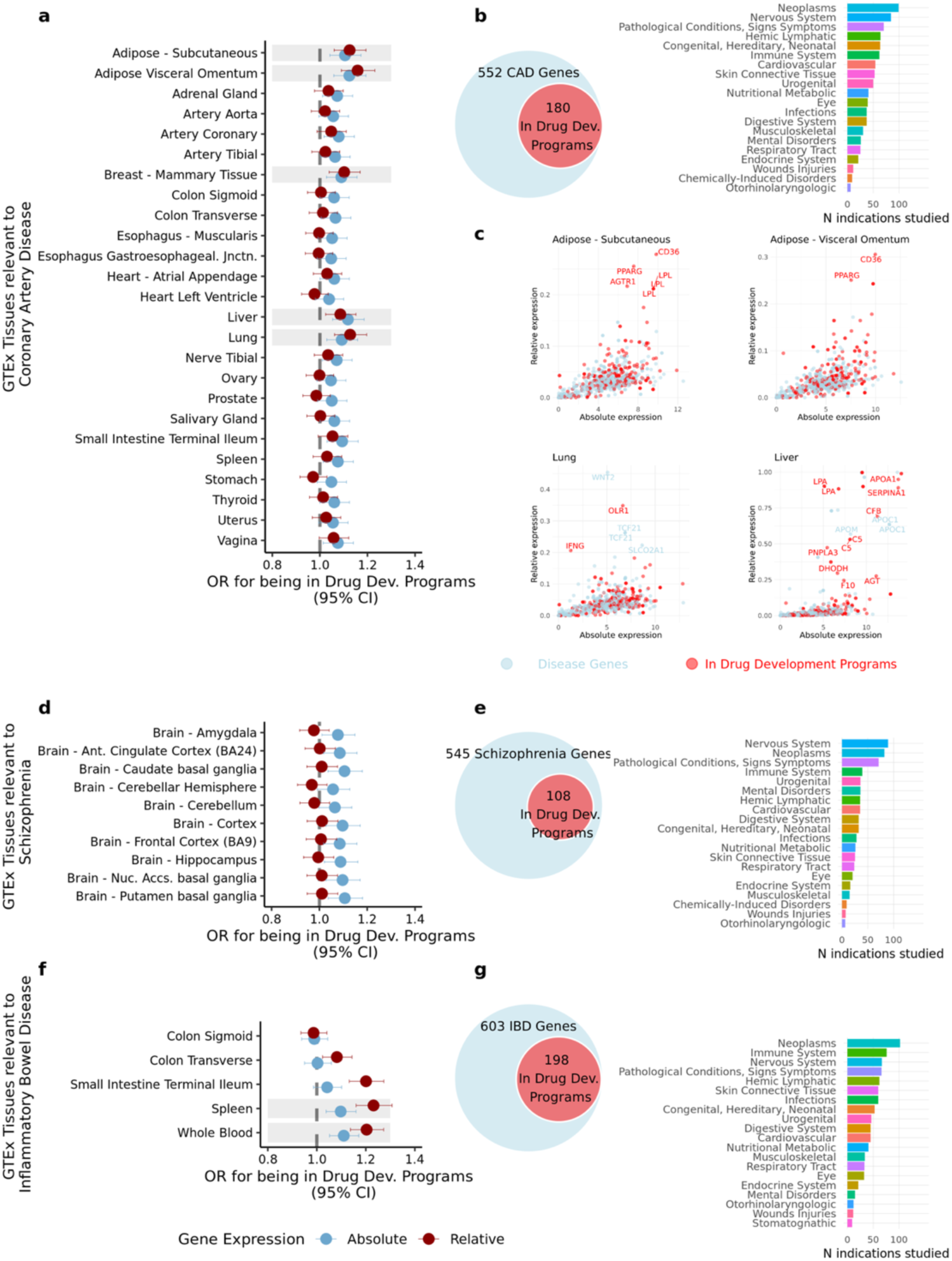
Impact of gene expression features on the inclusion of disease genes in drug target development. a, d, **f**, Forest plots displaying the odds ratios (OR) for absolute and relative gene expression affecting the likelihood of disease-associated genes being incorporated into drug development programs for coronary artery disease (CAD), schizophrenia, and inflammatory bowel disease (IBD). Only relevant GTEx tissues – defined as those in which disease genes are significantly more expressed than control genes (based on the GWAS to Gene Expression approach) – are included. Shaded areas indicate tissues where both absolute and relative expression have a significant OR. **b, e, g**, Bar plots showing the number of disease-associated genes already included in drug development programs and the MeSH categories of the indications for which drugs were studied. **c**, Scatter plots depicting the relationship between absolute and relative gene expression in CAD-associated genes. Shown are four tissues where both measures significantly increased the likelihood of CAD genes being incorporated into drug development programs. Genes highlighted in red correspond to those that have been included in drug development programs.

Figure 6 presents results for CAD, SCZ, and IBD. CAD genes with higher absolute and relative expression in subcutaneous adipose, visceral adipose (omentum), breast mammary tissue, liver, and lung are more likely to be included in drug development programs − often for indications unrelated to CAD (Figure 6a). We further investigated this association and find that, out of the 552 CAD genes, 180 were investigated as potential drug targets for 518 different indications, including cancers (neoplasms), nervous system disorders, cardiovascular diseases, and other pathological conditions (Figure 6b). Figure 6c highlights in red CAD genes included in drug development programs relative to their absolute and relative expression in adipose tissues, liver and lung.

While SCZ genes with high relative gene expression are not more likely to be included in drug development programs, those with higher *absolute expression* in key brain tissues are (Figure 6d). Of the 545 SCZ genes, 108 have entered drug development programs, primarily for indications related to nervous system disorders, cancer, immune system disorders, pathological conditions, and mental health disorders (Figure 6e).

IBD genes with higher relative expression in the small intestine (terminal ileum), blood and spleen are more likely to be included in drug development programs (Figure 6f and **Supplemental Table 19**). These results are driven by 198 out of 603 IBD genes that are already included in drug development programs. Drugs targeting these genes were investigated for 529 indications related to cancer, the immune system, the nervous system, pathological conditions, signs and symptoms, hemic-lymphatic diseases, and infections (Figure 6f).

## Discussion

In this study, we systematically investigated where disease genes, prioritised using GWAS results, are most expressed in the body, enabling insights into the initiation of disease and the pleiotropic effects of disease genes. Using different definitions for disease and control genes, three gene expression datasets at the tissue and cell type level, two measurements of gene expression, and alternative analytical strategies, we consistently find that disease genes are highly expressed in a broad range of tissues and cell types, including some without known links to disease. Higher expression of disease genes in several of those tissues is also associated with a greater OR for being included in drug development programs – often for indications unrelated to the original disease in which the genes were prioritised. Altogether, these findings could inform strategies for predicting side effects^43^ and repurposing drugs targeting disease genes.

While we use different analytical strategies to interrogate where disease genes act, a key novel aspect of our study is the *GWAS to Gene Expression* approach. Previous work has largely focused on analyses performed in the reverse direction – identifying tissues or cell types enriched for GWAS signals (i.e., the *Gene Expression to GWAS* approach) ^20,21,44^ – which we also use here to triangulate our results. However, the *GWAS to Gene Expression* approach that we introduce here is based on high-confidence causal disease genes, identified through state-of-the-art prioritisation methods applied to large-scale GWAS data. By focusing on genes most likely to be causal, this approach offers a more targeted framework for drug discovery than broader enrichment analyses. Our work also differs from other approaches that combine GWAS and functional genomic data to make inference about disease aetiology. For example, Transcriptome-Wide Association Studies (TWAS) integrate GWAS findings with expression quantitative loci (eQTLs) to investigate how genetic variants influence gene expression^30^. TWAS relies on the availability of eQTL data and on genes with highly heritable gene expression^45^. In contrast, our approach provides a gene expression profile of disease-associated genes regardless of eQTL data availability and gene heritability, broadening the scope for combining GWAS signal and gene expression.

We first focus our study on tissue-level analysis because extensive prior knowledge of disease-tissue associations provides a “ground truth” and, thus, informs the benchmarking of our approach. Next, we apply our approach to Tabula Sapiens and ARCHS4 datasets, which offer insights not easily discerned at the tissue level. For example, the results that we observe between AD and IBD in spleen, blood and lung are likely driven by the high fraction of macrophages and other innate immune system cells present within those tissues. However, cell-type level datasets also have limitations: in the case of ARCHS4, the representation of cell types and tissue regions is less systematic than in GTEx. Furthermore, ARCHS4’s heterogeneity may have reduced power to detect associations in other diseases despite correction of batch effects. While *Tabula Sapiens*^25^ may provide a more systematic multiorgan dataset at the cellular level, the scale of this dataset is smaller (they measured single-cell RNA-sequence data for a total of 15 individuals, respectively, in contrast to e.g. GTEx, which assessed more than 700 individuals). Moreover, some tissues of interest were not available here, such as brain tissues to interrogate cell types related to AD, SCZ, BP or ADHD.

There are two key considerations when interpreting our results and implementing our pipeline. First, no gene prioritisation method is perfect, and therefore some genes categorised as ‘disease genes’ may not, in fact, affect disease. Moreover, prioritisation methods that used functional genomics datasets may be more powered, but they may provide circular results, especially when using functional data from tissues hypothesised to be implicated in the disease. This could be particularly relevant for the analyses of cancer traits, since prioritisation approaches often rely exclusively on the specific cancer tissue RNA-seq dataset. Second, this study mainly used expression datasets that included tissues and individuals considered to be healthy. The gene expression profiles represent a healthy state and cannot be used to explore the dynamics of gene expression during disease states. However, before studying the dynamics of gene expression between cases and controls, we argue that it is important to understand the tissues and cell types in which disease genes show high expression and specificity.

In conclusion, we present a comprehensive expression profiling of genes identified as high-confidence disease genes. We identify known and novel tissues and cell types that could have causal links to disease, factors that contribute to variability in the gene expression of disease genes, and explore the potential relevance of gene expression to the inclusion of disease genes in drug development programs. All together, we hope that this work opens new avenues for guiding experimental follow-up and informs future drug development efforts targeting genes with support from GWAS data, ultimately advancing our understanding of disease mechanisms and response to treatment.

## Methods

### RNA-seq datasets

#### GTEx dataset

Gene expression measurements were obtained for 50 tissues from the GTEx project^24^ version 8. Median gene TPMs for each tissue were downloaded from https://gtexportal.org/home/datasets. Standard RNA-seq processing steps were applied to the dataset as follows: (1) we filtered out all non-protein-coding genes and genes not expressed in any tissue; (2) we removed the tissues with less than 100 samples, cancer or cell related tissue types (i.e. EBV-transformed lymphocytes and Leukemia cell lines); (3) we scaled the expression of each tissue such that the total is 10^6^ TPM. 45 tissues remained after quality control.

#### Sex-stratified analyses in the GTEx dataset

In tissues for which we wanted to test whether the disease-associated genes present higher absolute or relative expression in men and women specifically (i.e. T2D), we extracted the GTEx expression dataset, and for each tissue, the median gene expression of all genes was calculated for women and men separately. For each sex, we then created absolute and relative expression datasets (where columns represent tissues, and rows represent genes). To prepare sex-specific inputs for the MAGMA analyses, we generated GMT files for men and women separately, which contain the gene sets composed of the top decile of absolute and relative gene expression for each sex. T-tests, Anderson-darling tests, and MAGMA analyses were also performed separately for men and women.

#### ARCHS4 dataset

Gene expression across cell types and tissue regions were extracted from the ARCHS4^25^ resource, which provides access to uniformly processed gene counts from human RNA-seq experiments stored in the Gene Expression Omnibus (GEO) and Sequence Read Archive (SRA). Using the ARCHS4 web browser (https://maayanlab.cloud/archs4/), we systematically identified all the cell types available in the metadata search menu. For some disease-relevant cell types like microglia in AD, we entered the cell type name directly into the metadata search bar. ARCHS4 generates R scripts listing the samples related to each cell type, to facilitate their extraction from the main repository −a HDF5 file named “human_gene_v2.2.h5” available for download at https://maayanlab.cloud/archs4/download.html (Downloaded version date: 5-30-2023).

The quality control of ARCHS4 datasets and *per-gene* TPM calculation was as follows: Only cell types with more than 100 samples across all experiments available in ARCHS4 were included in our analyses. Upon obtaining the counts expression matrix for each cell type, we performed quantile normalisation of samples using the function ‘*normalise.quantiles’* available on the R package (”preprocessCore”). Quantile normalisation was performed on raw counts. Given that samples from a specified cell type may originate from multiple experiments with slightly different conditions, we (1) excluded experiments containing less than 10 samples, and (2) adjusted for batch effects using the package ComBat_seq^46^, which is an improved version of the popular ComBat^47^. Unlike its predecessor ComBat (designed for microarray data), ComBat_seq is tailored for RNA-Seq studies and it does not assume a normal distribution of gene expression data.

After batch correction, median TPM values for each gene were calculated in each cell type using the formula:

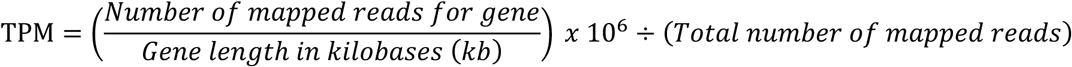

where ‘Gene lengths in kilobases’ were calculated using the genomic coordinates indicated in a GTF file (built GRCh38, version 106), downloaded from ENSEMBL (https://ftp.ensembl.org/pub/release-106/gtf/homo_sapiens/Homo_sapiens.GRCh38.106.gtf.gz).

#### Tabula Sapiens datasets

scRNA-seq datasets were obtained from the *Tabula Sapiens* figshare (https://figshare.com/articles/dataset/Tabula_Sapiens_release_1_0/14267219). These datasets, initially in ‘.h5ad’ format, contained gene counts for each cell and metadata, and were converted into Seurat objects and then into ‘*SingleCellExperiment*’ objects to ensure the input data files were in a similar format as the ones used for ARCHS4 and GTEx.

Data quality processing was performed: cells with zero counts across all genes were removed, and outlier cells with an extreme number of detected genes were excluded. Pseudobulk data was then generated using the *aggregateToPseudoBulk* function from the *dreamlet* R package to aggregate expression counts across cell types, according to the free cell type annotation included in the original datasets.

#### Calculating ‘Absolute gene expression ‘and ‘Relative gene expression’ values

Since the expression patterns of protein-coding genes tend to follow a negative binomial distribution, we calculated *absolute levels of gene expression* by taking the Log_2_ of the median TPM+1 values. To calculate *relative gene expression*, we divided the *absolute levels of gene expression* of each gene by its total expression across tissues. The resulting relative gene expression ranged from 0 (gene is not expressed) to 1 (gene is exclusively expressed in this tissue). The Log_2_ of the absolute and relative expression measures were used in subsequent analyses.

To calculate absolute and relative gene expression values for each gene in the *Tabula Sapiens* dataset, the total expression counts for each cell type was obtained, and normalisation was performed using the *calcNormFactors* function from the *edgeR* R package. Absolute gene expression values were calculated as the total number of counts per million for each gene in each cell type (after normalisation). Data was organised in data tables where each row represented a gene, and each column represented a cell type. For relative expression of a gene in a cell type, the *cellTypeSpecificity* function – which calculates the number of counts of a gene in a cell type divided by the total number of counts across cell types in that tissue – was utilised after applying the same normalisation procedure as for absolute expression.

### GWAS to Gene Expression

#### Definition of disease-associated genes inferred from GWAS results

Three different types of gene lists were inferred for each disease using GWAS results: nearest-to-hit genes, fine-mapped genes, and PoPS genes. For all the gene definitions, we obtained each gene ENSEMBL IDs using a GTF file (built GRCh37, version 75), downloaded from ENSEMBL https://ftp.ensembl.org/pub/release-75/gtf/homo_sapiens/Homo_sapiens.GRCh37.75.gtf.gz). The list of prioritised genes for each approach and disease can be found in **Supplemental Tables 1-3.**

#### Definition of ‘nearest-to-hit’ genes

To find the genes closes to the GWAS hit, we obtained publicly available GWAS summary statistics for the diseases investigated. We performed clumping using PLINK 1.9^48^ and individual level genotype data from the 1000 Genomes Project^49^ as a reference linkage-disequilibrium (LD) panel. During clumping, variants with *P-* values ≤-s 5×10^-8^ were retained, and variants within a 250 Kbp window correlated ≥ 0.5 with the index variant or variants with *P-*value ≥ 0.01 were removed. For each clump, the nearest protein-coding gene to the index variant was identified. We used the GTF file (built GRCh37, version 75) to extract the gene start and gene end coordinates of each protein-coding gene. Information about the GWAS used, number of clumped variants, genes identified and distance between variant and nearest gene is included in **Table 1**.

#### Definition of ‘fine-mapped genes’

We acquired gene lists that previously fine-mapped GWAS for CAD^3^, SCZ^4^, IBD^5^, AD^53^, BD^7^, ADHD^8^, T2D^9^ and Vitamin D^54^. Most gene lists were constructed using a combination of statistical fine-mapping, transcriptome association studies, and mendelian randomization.

For CAD, the integration of eight gene prioritisation predictors enabled the identification of 220 likely causal genes^3^. For SCZ, statistical fine-mapping was integrated with summary Mendelian randomization and Hi-C interaction data to obtain a list of 120 prioritised genes^4^. For IBD, we extracted the list of genes linked to variants fine-mapped, available in the Supplemental material of the study conducted by Huang and colleagues^5^. For AD, we used a review of that reported a list of genes prioritised via fine mapping of GWAS in two previous studies^10,11^. The list is available in https://github.com/sjfandrews/ADGenetics/blob/main/results/adgwas_loci.csv. For BP, fine-mapping of the GWA signals was performed and seven complementary approaches were used to prioritise 47 credible genes that were mapped to loci by at least three of the seven approaches^7^. For ADHD, fine-mapping of the most recent ADHD GWAS^8^ identified sets of credible variants for each risk locus. Credible sets were subsequently linked to genes based on genomic position, information about eQTLs, and chromatin interaction mapping in human brain tissue as implemented in FUMA. For T2D, we used a gene list containing the nearest gene of the results of a fine-mapping approach used in 380 independent association signals^9^. For Vitamin D, we extracted a list of genes published by *Manousaki* and colleagues^54^, who prioritised genes using the DEPICT method^28^ on a GWAS of serum 25 hydroxyvitamin D.

#### Definition of ‘PoPS genes’

The PoPS method^31^ prioritises disease-associated genes by integrating gene-level z-scores from MAGMA^22^, single-cell gene expression data, biological pathways, and predicted protein-protein interaction networks. The original PoPS publication suggests combining PoPS scores with location information would provide a list of high-confidence genes. However, the combined PoPS+location approach leads to a low recall (it detects very few genes for each disease). Therefore, we used the top 1% PoPS, because it results in a list of 184 prioritised genes per disease. This number of genes is similar to the number of fine-mapped and nearest-to-hit genes, reducing differences in power due to the number of genes assessed. The choice of the parameters top 1% of PoPS scores is further discussed in the **Supplemental Material**; our results are robust across thresholds (**Additional File 1, Supplemental Figure 3**). Full PoPS scores are available at: https://www.finucanelab.org/data.

### Definition of control genes

In this study, three control groups are defined. The control group used for our primary analyses was defined by all protein-coding genes. Additionally, two additional control groups were tested, using genes associated with other diseases, identified by the Open Targets initiative.

#### Definition of ‘control genes’ in our primary analysis

We defined the control group as all protein-coding genes expressed in the tissue or cell type being analysed. These results form the basis of our main findings (e.g., Figures 2, 3, and 4). To identify these genes, we used the publicly available GTF file (built GRCh38, version 106), downloaded from ENSEMBL (https://ftp.ensembl.org/pub/release-106/gtf/homo_sapiens/Homo_sapiens.GRCh38.106.gtf.gz). Using this file, we selected genes located on autosomes with the biotype designation “protein-coding.” Any protein-coding genes not included in the list of disease-associated genes were classified as part of the control group.

#### Definition of ‘control genes’ groups, utilising genes associated with other diseases

To generate a list of control genes associated with multiple traits and diseases, we extracted two lists of genes from the Open Targets resource^55^. The first group uses the *Open Targets ‘Gold Standards’*. The second group uses a list of genes prioritised via *Open Targets Genetics evidence,* using a machine learning method^56^ that calculates a disease-specific score to prioritise genes.

Open Targets Gold standards: This list of genes represents a repository of >400 published GWAS loci for which there is high confidence in the gene functionally implicated. The list of gold standard genes was downloaded from https://github.com/opentargets/genetics-gold-standards/blob/master/gold_standards/processed/gwas_gold_standards.191108.tsv. The final set of genes was composed by 519 protein-coding genes from 284 traits were used as gold standard control gene list. The traits with the largest number of genes were 2 diabetes (44 genes), breast carcinoma (43 genes) and prostate carcinoma (23 genes).

Open Targets Genetics evidence: This list of genes was extracted from the results of a machine-learning method used to identify the most likely causal genes^56^. This method integrates the results of 1) fine-mapping credible set analysis, 2) functional genomics data such as pathogenicity prediction, colocalization with molecular QTLs, genomic distance and chromatin interaction data to generate predictive features. The machine-learning model is supervised using the gold-standard positive GWAS loci, and a score is computed for each gene (named Locus to gene (L2G) score). The L2G score is calibrated so that a gene’s score indicates the fraction of genes at or above the score that would be expected to be true positives. Thus, we selected genes with a score >= 0.8, which assumes that 80% of the genes associated with a trait or disease in our list are causal.

Data was downloaded from the publicly available website https://platform.opentargets.org/downloads/data, section “Target - Disease evidence / Integrated list of target - disease evidence from all data sources” (version 23/09), which provides several directories with different evidence sources for the target-disease associations. However, only the ones indicating ‘genetics evidence’ were used in our analyses. From those, 3,862 protein-coding genes from 1,582 traits had L2G scores >= 0.8. The traits with the largest number of genes were height (580 genes), blood protein measurement (359 genes), and heel bone mineral density (286 genes).

### Statistical analyses to compare disease-associated genes vs control genes

The *t-*test is an inferential statistic used to evaluate whether the means of two independent samples are significantly different. Here, we run one-side *t-*tests in R, testing the null hypothesis that the expression of disease-associated genes is higher than those of the control group. *t-* tests assume that the sample means are normally distributed. Since gene expression follows a negative binomial distribution, we normalised the gene expression values by taking the Log_2_ of the median TPM+1 before applying the *t-*tests.

The Anderson-Darling^34^ is a non-parametric test to evaluate whether the gene expression of disease-associated genes originates from the same distribution than the control group of genes. It tests the null hypothesis that both groups were drawn from populations with identical distributions. The Anderson-Darling test is similar to other tests assessing differences between empirical distributions (such as the two-sample Kolmogorov-Smirnov test^57^), but it is more sensitive to differences in the tails of the distribution.

For the Tabula Sapiens dataset, *P*-values to compare disease-associated genes vs control genes were calculated using a permutation approach (10,000 permutations). We calculate empirical *P*-values here to account for the small sample size of the dataset (up to 15 individuals, although typically only 2 individuals were used to extract scRNA-seq measurements for each tissue). This method avoids bias in cases where cell types are obtained from cells derived from the same individual, ensuring that results are not affected by violated assumptions of independence –which would invalidate a t-test.

### Gene Expression to GWAS

#### Definition of genes in the top decile of expression

For each tissue, we defined to gene-sets that are in the top decile of gene expression: one gene-set is composed by the 10% of genes with the highest *absolute* gene expression. The second gene-set is composed by the 10% of genes with the highest *relative* gene expression. To obtain these gene-sets, we first classified all protein-coding genes into 11 quantiles. In this classification, the 1st quantile is composed by genes without expression in a specific tissue, whereas the 11th quantile encompasses the genes with the highest expression values. We then grouped genes from the top quantile and tested their GWAS enrichment using MAGMA and the 1000Genomes as a reference panel. We expanded the gene coordinates by adding a 35 kb window upstream and a 10kb window downstream of the gene. The Major Histocompatibility Complex (MHC) region was excluded from the analyses due to their long-range LD.

#### Gene set enrichment analyses with MAGMA

MAGMA^22^ is a software designed for gene-set enrichment analysis using GWAS data. It provides enrichment results at the gene-level and at the gene-set level. In gene-level analyses, MAGMA employs GWAS *P-*values to compute gene test statistics, accounting for LD structure via a reference dataset. For gene-set analyses, gene-level association stats are transformed into Z-scores, reflecting the strength of gene-phenotype associations. MAGMA uses a competitive gene-set test formula: Z = β₀ + Iβₚ + Cβₖ + ɛ where I is an indicator (1 if a gene is in gene-set p, 0 if not), and C is a covariate matrix. The resulting *P-*value originates from a test on coefficient βₚ, evaluating if the phenotype shows a stronger association with genes included in the gene-set of interest versus other genes.

In our analyses, SNPs from GWAS data were mapped to genes based on genomic coordinates defined in a GTF file (Genome build GRCh37). Gene boundaries were extended 35kb and 10Kb around the 5’ and 3’ gene boundaries, respectively. The reference dataset used to account for LD was the genotype data from the 1000 Genomes Project.

### PubMed search

To determine which tissues are associated with specific diseases based on previous knowledge, we interrogated how often a combination of tissue and disease terms appeared together in published articles found on PubMed. To count the PubMed occurrences of a tissue being mentioned in relation to a disease, we used the Python library *Beautiful Soup*^58^, taking the queries defined above as input. The script performs the following tasks: it generates combinations of tissue-disease pairs, constructs search queries, sends requests to the PubMed website based on these queries, and subsequently extracts the number of search results from the webpage. The resulting count shows how frequently the tissue-disease pair appears in the body of literature. Two types of PubMed search analyses we conducted based on the type of query constructed: one using Medical Subject Headings (MeSH) Terminology, and another one using keywords.

#### Search queries utilising MeSH Terminology

The Medical Subject Headings (MeSH) thesaurus is a curated collection of terms established by the National Library of Medicine. MeSH terms are valuable in recognizing content that uses different words but refers to the same concept, enhancing the accuracy and consistency of the literature search process. Leveraging MeSH terminology, we prioritised the list of 45 tissues from the GTEx dataset based on their frequency of occurrence within MeSH terms connected to scientific articles. For each pairing of tissue and disease, a search query in the format *‘<TISSUE name> [Mesh] AND <DISEASE name> [Mesh]’* was used.

#### Search queries utilising Keyword-based Literature Search

The list of tissues and cell types were ranked based on their citation frequency within the titles or abstracts of relevant scientific articles. The construction of search queries followed the format ‘*<TISSUE name> [Title/Abstract] AND <DISEASE name> [Title/Abstract]’* for each unique tissue-disease pair.

### Calculating predictors of disease-associated gene expression

To uncover the key contributors to the variability in gene expression among disease-associated genes, we performed variance partition analyses using the R package ‘*variancePartition*’^38^. This package assesses drivers of variation for each gene by fitting a linear fixed model to quantify the contribution of tissues, individuals, technical variables etc. in gene expression.

We calculated the variance partition for each disease-associated and control gene. We used as predictors uncorrelated variables (r^2^ < 0.75) that explained the largest proportion of variance in gene expression, as calculated by the Canonical Correlation Analysis in the *variancePartition* package and reported in the original *variancePartition* publication^38^ (which also used the GTEx dataset). The variance partition analysis results in a data table where each row is a gene, and each column is the predictor variable included in the model. The results show, for each gene, the percentage of variance explained for each predictor.

### Assessing the impact of gene expression on the inclusion of disease genes in drug development programs

To evaluate whether absolute and relative gene expression influence the likelihood of disease genes being considered as drug targets, we assessed whether higher absolute and relative expression values across GTEx tissues increase the odds of disease genes being included in drug development programs.

We extracted drug development data for all disease genes from the Citeline Pharmaprojects database, as curated by Minikel et al.^12^. This dataset includes monotherapy programs initiated since 2000, annotated with the human gene target for each drug under study and for each disease indications the program was developed. The table was downloaded from the study’s Github repository (https://github.com/ericminikel/genetic_support/blob/main/data/pp.tsv).

For each disease investigated, we combined the three lists of disease-associated genes used for the “GWAS to gene expression” approach: (i) nearest-to-GWAS hit genes, (ii) fine-mapped genes, and (iii) PoPS-prioritised genes (top 1% PoPS threshold). Disease genes absolute and relative gene expression for each GTEx relevant tissue were extracted. Relevant tissues were defined using the same criteria as in the sensitivity analysis that excluded genes highly expressed in disease-relevant tissues (**Additional File 1, Supplemental Material**). Specifically, tissues were considered relevant if they showed significantly higher expression in all statistical tests performed. A Bonferroni-corrected significance threshold was set at 0.05 / (45 x 3 x 2) to account for 45 tissues, 3 gene prioritisation approaches, and 2 statistical tests (t-test and Anderson-Darling test). Tissues where both tests yielded *P*-values < threshold were selected. Expression values were classified into quantiles (10 and 100 quantiles were tested, leading consistent results) to mitigate the effect of genes with extreme expression values inflating the OR.

To assess the impact of gene expression on the inclusion of disease genes in drug development programs, we performed logistic regression separately for each gene expression feature (absolute or relative), for each disease and for each relevant tissue. The dependent variable was a binary indicator, where each disease gene was assigned 1 if included in a drug development program and 0 otherwise. The independent variable was either absolute or relative gene expression quantiles of that gene in the relevant tissue. OR and 95% Confidence intervals were calculated from the output of the logistic regression.

To categorize the indications for which drug targets had been investigated into broader groups (as shown in Figure 6, panels b, e, and g), we extracted the MeSH IDs associated with all indications from the Minikel dataset and mapped them to their ancestor categories in the MeSH ontology tree. To retrieve the ontology hierarchy for each MeSH ID, we queried the MeSH Resource Description Framework (RDF) dataset using SPARQL.

## Supporting information

Additional File 2 - Main Analyses

Additional File 3 - Cancer Analyses

## Data Availability

All data produced are available online and in the supplemental materials. The scripts used in the current study are available at https://gitlab.com/JuditGG/GeneExpressionLandscape

https://juditgg.shinyapps.io/diseasegenes/

## Code availability

We have released an open-source implementation of our approach, along with all derived gene lists from publicly available data (https://gitlab.com/JuditGG/GeneExpressionLandscape). This pipeline provides a flexible framework for analysing prioritised gene sets across diseases and can be adapted for future studies.

## Acknowledgements

This work was supported by a grant from the National Institutes of Health (R01MH122866) to PFO, by a 2022 NARSAD Young Investigator Grant (Number 30749) by the Brain & Behavior Research Foundation to JGG, and a grant from the National Human Genome Research Institute (1K99HG013547-01) to JGG.

Additionally, this work was supported in part through the computational and data resources and staff expertise provided by Scientific Computing and Data at the Icahn School of Medicine at Mount Sinai and supported by the Clinical and Translational Science Awards (CTSA) grant UL1TR004419 from the National Center for Advancing Translational Sciences. Research reported in this publication was also supported by the Office of Research Infrastructure of the National Institutes of Health under award number S10OD026880 and S10OD030463.

The content is solely the responsibility of the authors and does not necessarily represent the official views of the National Institutes of Health.

We thank the participants of the ARCHS4, Tabula Sapiens and the GTEx projects and the scientists that shared their results involved in the construction of these resources.

## Author contributions

JGG: Conceptualization, Funding Acquisition, Data Curation, Formal Analysis, Investigation, Software, Validation, Supervision, Visualization, Writing – Original Draft Preparation, Writing – Review & Editing. AC: Investigation, Software, Validation. SGG: Investigation. LL: Writing – Review & Editing, Data Curation. PFO: Conceptualization, Funding Acquisition, Formal Analysis, Supervision, Writing – Review & Editing.

