## Additional File 1 for "The Gene Expression Landscape of Disease Genes"

#### Table of Contents

Sex-stratified analyses for Type 2 Diabetes 2

Anderson-Darling Test Results 5

Considerations and Analysis for PoPS Threshold Selection 7

GWAS to Gene Expression Results Using Open Target Genes as Control Groups 10

Sensitivity analysis where highly expressed genes are removed 18

The Gene Expression Landscape of Disease Genes in the ARCHS4 database 20

Supplemental References 37

#### Sex-stratified analyses for Type 2 Diabetes

Since results show T2D genes in breast present higher expression than control genes, we performed sex-stratified analyses to see whether results were driven by one of the sexes. We repeated our analysis and compared the gene expression of disease *vs* control genes in men and women separately (**Methods**).

For T2D, disease associated genes are more expressed in breast for both sexes (**Supplemental Figure 1, panels a and b**), although *P*-values were slightly lower for men (*P*-values for Rel. and Abs. Expression < $10^{-6}$) than for women (*P*-values for Rel. and Abs. Expression < $10^{-4}$). Significant results were also observed for adipose tissues (*P*-values range: $10^{-3}$ to $10^{-13}$), pituitary in the case of women (PoPS *P*-value=0.001), and testis in the case of men (PoPS *P*-value=$5.68 \times 10^{-12}$). Significant results were observed only for the PoPS gene list, and had smaller *P*-values in the Anderson-Darling test than in the *t*-tests (**Supplemental Table 20**).

To investigate whether specific genes are driving the T2D results in a sex-specific manner, we examined the expression of individual genes for each gene list (nearest-to-hit, Fine-mapped, PoPS), even though results were only significant for PoPS. Across all tissues with significant P-values, Thymosin Beta 10 (TMSB10), Cluster of Differentiation 74 (CD74), and Complement Factor D gene (CFD) were PoPS genes with high expression values (**Supplemental Figure 1, Panel c**).

TMSB10, a gene involved in the regulation of actin polymerization and cytoskeletal organization, has been implicated in various cancers [1–4], including pancreatic cancer [5,6], which is of particular relevance given the epidemiological link between T2D and an increased risk of pancreatic cancer. Individuals with T2D have been shown to have a two-fold higher risk of developing pancreatic cancer [7,8], highlighting the importance of understanding TMSB10's role in both conditions. Additionally, both CD74, a glycoprotein essential for immune response regulation, and CFD, a protein involved in the alternative complement pathway, play key roles in chronic inflammation[9,10], which is a hallmark of T2D[11]. Since the inflammatory processes in T2D are closely linked to insulin resistance, and these genes may be pivotal in modulating that response, high expression of these genes could be relevant in T2D pathogenesis and they could serve as promising candidates for further investigation.

Sex-stratified analyses for Type 2 Diabetes

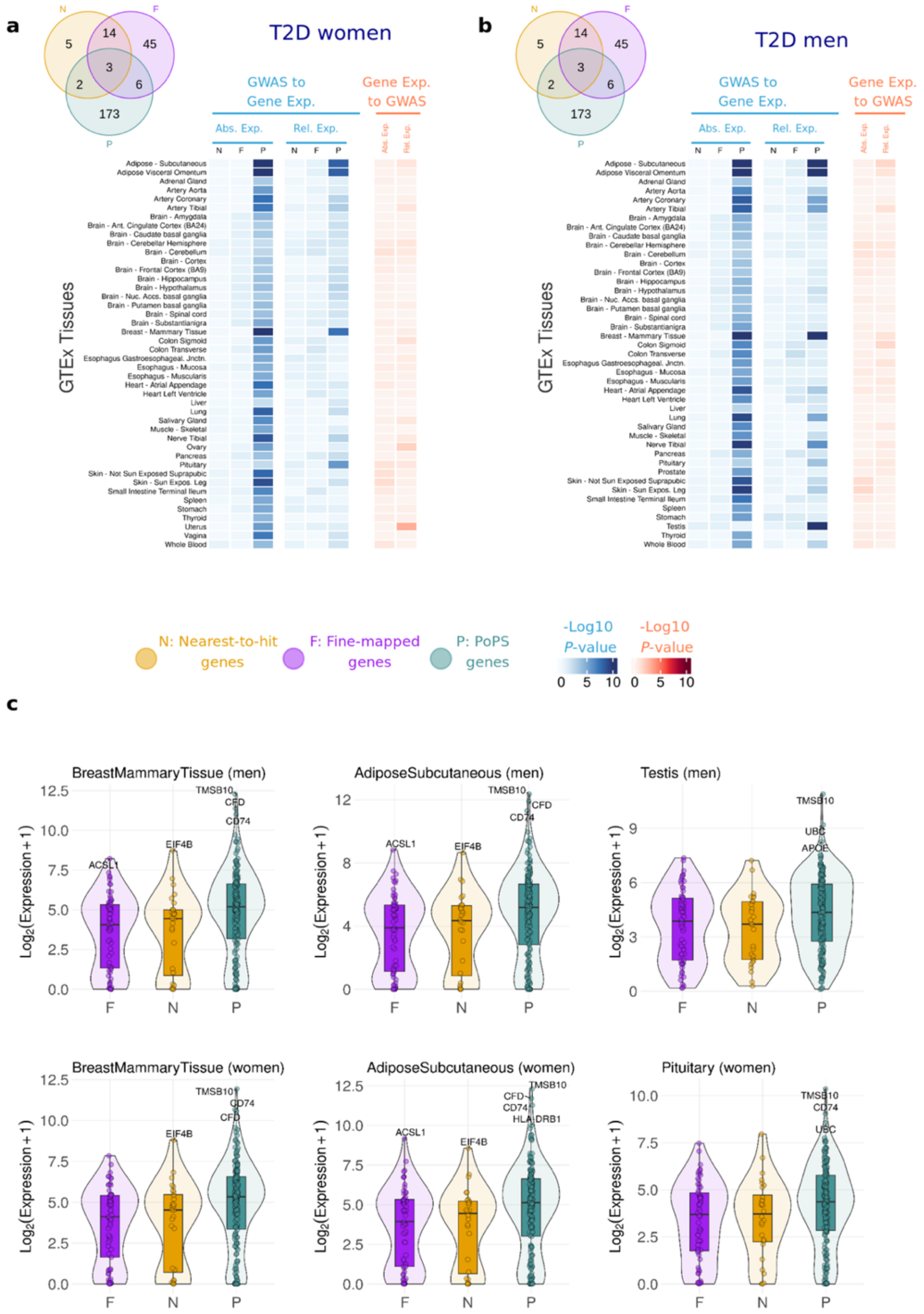


**Supplemental Figure 1: Sex-stratified analyses for T2D.** Heatmap showing results of the association between gene expression in each GTEx tissue and Type 2 Diabetes for women (a) and men (b). In blue, results showing the $Log_{10}$ *P*-value for a one-side *t*-tests, testing the null hypothesis that disease-associated genes are not more expressed than other protein-coding genes expressed in that tissue. In red, results showing the $Log_{10}$ *P*-value for enrichment of GWAS signal across the set of genes with highest absolute and relative expression for each tissue. Abs. Exp, Absolute expression; Rel. Relative expression; c) Gene expression levels of disease genes for Breast Mammary tissue, Adipose subcutaneous and Men and Pituitary. Results are presented for each sex separately, indicated in brackets. F, Fine-mapped genes; N, Nearest-to-hit genes, P, Polygenic Priority Scores genes.

Anderson-Darling test results

### Anderson-Darling Test Results

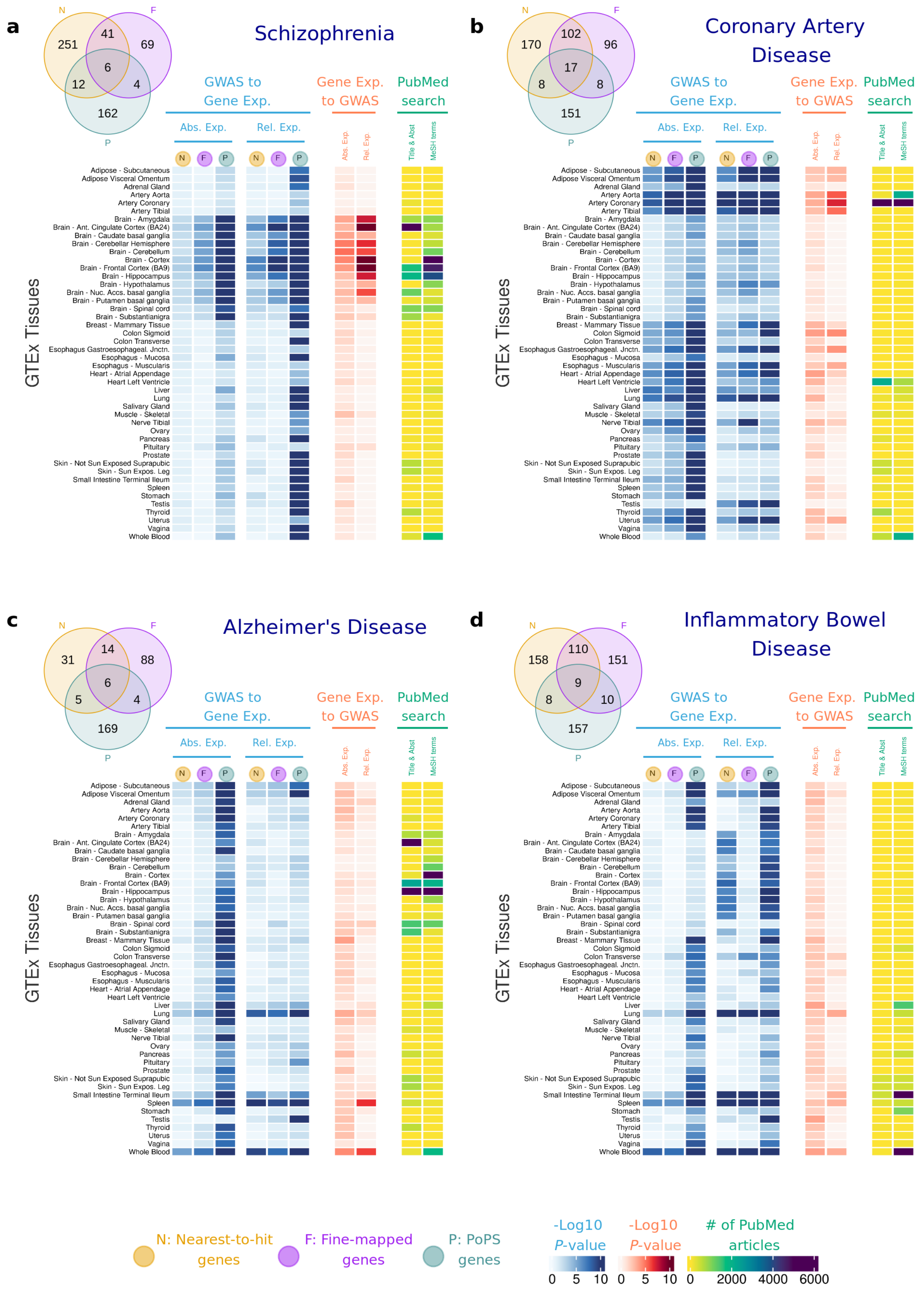

Anderson-Darling test results

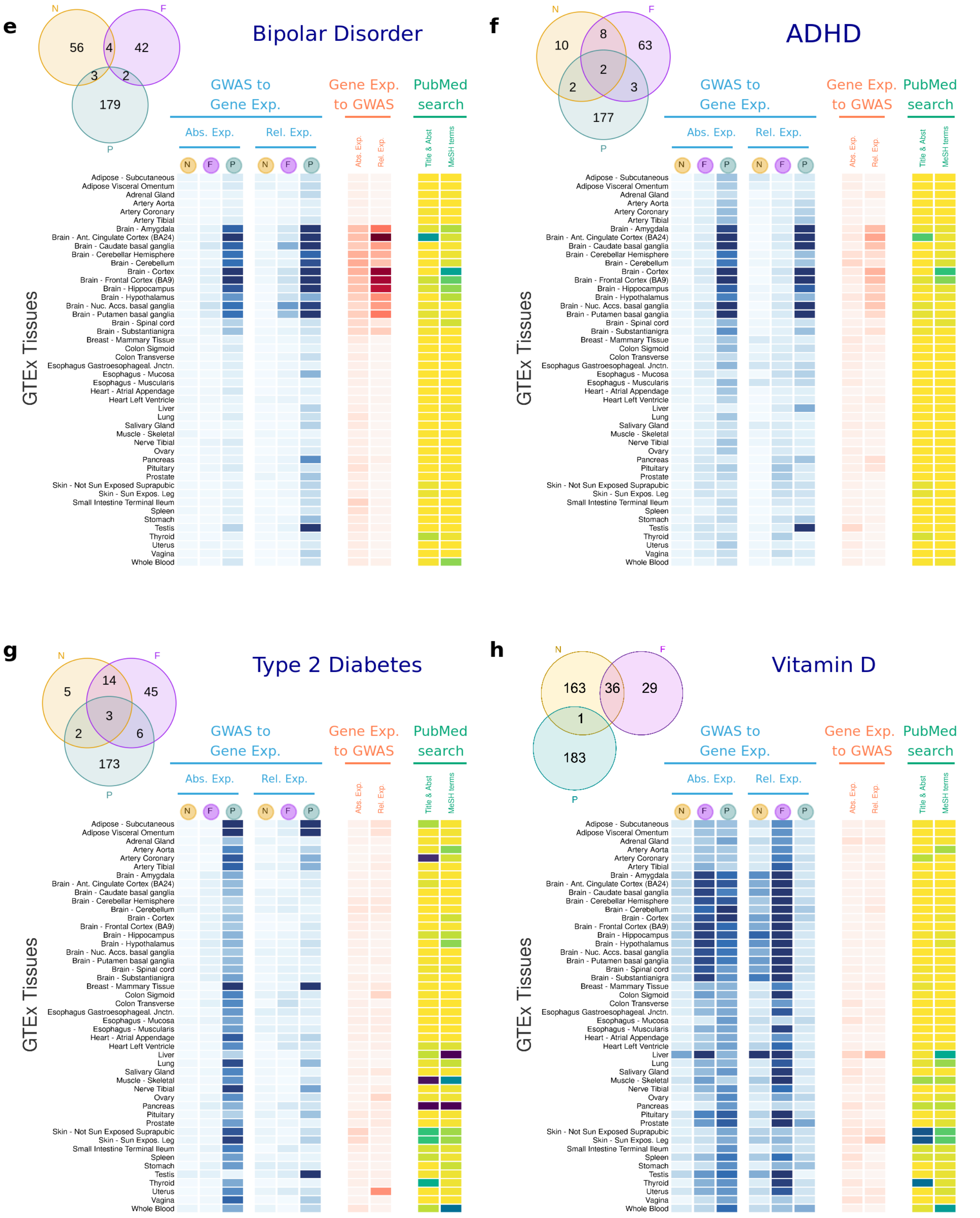


**Supplemental Figure 2:** Heatmap showing results of the association between gene expression in each GTEx tissue and **a**, Schizophrenia; **b**, Alzheimer's Disease; **c**, Coronary Artery Disease; **d**, Inflammatory Bowel Disease; **e**, Bipolar Disorder; **f**, ADHD; **g,** Type 2 Diabetes; **h**, Vitamin D. In blue, results showing the P-value of the **Anderson-Darling test**, (as opposed to the One side T-test shown in Main Figure 2). In red, results showing the P-value for enrichment of GWAS signal across the set of genes with highest and most specific expression for each tissue. In yellow, results for the Literature Search using PubMed. F, Fine-mapped genes; N, Nearest-to-hit genes, P, Polygenic Priority Scores genes; TiAb, Title and Abstract are used in the PubMed Search; MeSH, MeSH terms are used in the PubMed Search.

#### Considerations and Analysis for PoPS Threshold Selection

We conducted sensitivity analyses using varying PoPS score thresholds across the eight major diseases and three cancers included in this study, all GTEx tissues, and both absolute and relative gene expression. Specifically, we performed *t*-tests comparing the means of disease-associated genes –defined by the top 0.5% (92 genes), 1% (184 genes), and 5% (919 genes) of PoPS scores –against all other protein-coding genes as controls.

Our results indicate that overall, findings are robust to the choice of PoPS thresholds (**Supplemental Fig. 3**). Disease-associated genes consistently exhibit higher expression levels in tissues with known disease relevance (e.g., brain tissues for psychiatric disorders such as ADHD, schizophrenia, and bipolar disorder; liver for Vitamin D metabolism) as well as in less expected tissue-disease associations (e.g., blood, lung, adipose, and spleen for inflammatory bowel disease and Alzheimer's disease).

While overall the results are robust to the choice of the PoPS top percentage of genes used, these analyses also reveal differences between absolute and relative expression across PoPS thresholds:

- Relative gene expression results remain consistent across thresholds, with the lowest P-values observed at the 5% threshold.
- Absolute gene expression results suggest that more relaxed PoPS thresholds (e.g., 5%) increase gene expression differences across many tissues. We argue that this could inflate the rate of type I errors in our analyses. For example, in psychiatric disorders, the 5% threshold identifies significant expression in most tissues, whereas the 1% and 0.5% thresholds show significance primarily in brain tissues, aligning more closely with prior disease knowledge. As a result, the use of more stringent thresholds (0.5% or 1%) seems more appropriate for analyses comparing absolute gene expression.

Based on these results, we propose the 1% PoPS threshold is the most appropriate for the following reasons: (1) Relative expression results are stable across thresholds. (2) Absolute expression at the 1% threshold captures significant disease-control differences while minimizing type I errors, and (3) Gene set size at the 1% threshold (184 genes) is comparable to fine-mapped gene lists for many diseases (Supplemental **Table 2**), ensuring similar statistical power.

Considerations and Analysis for PoPS Threshold Selection

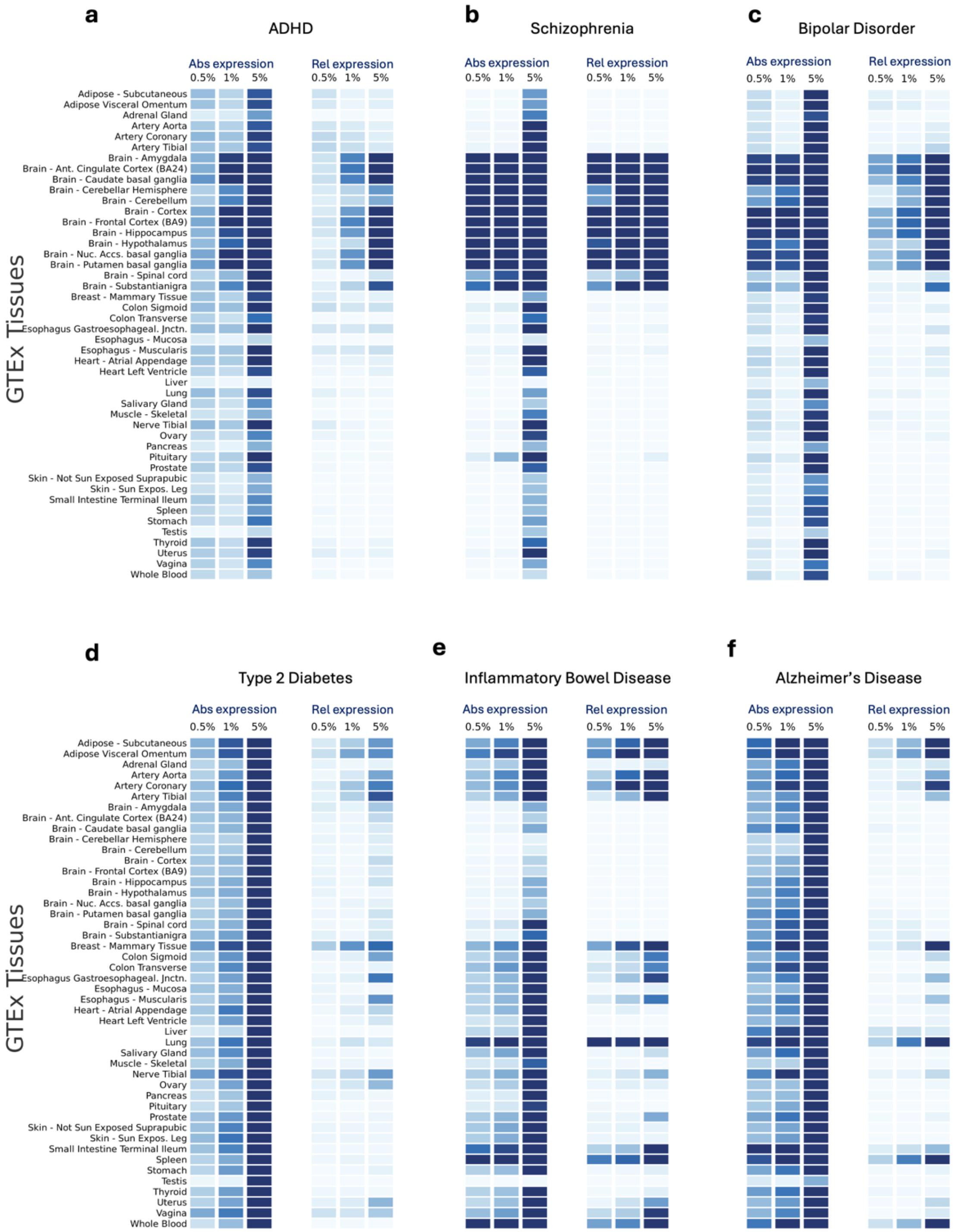

### Considerations and Analysis for PoPS Threshold Selection

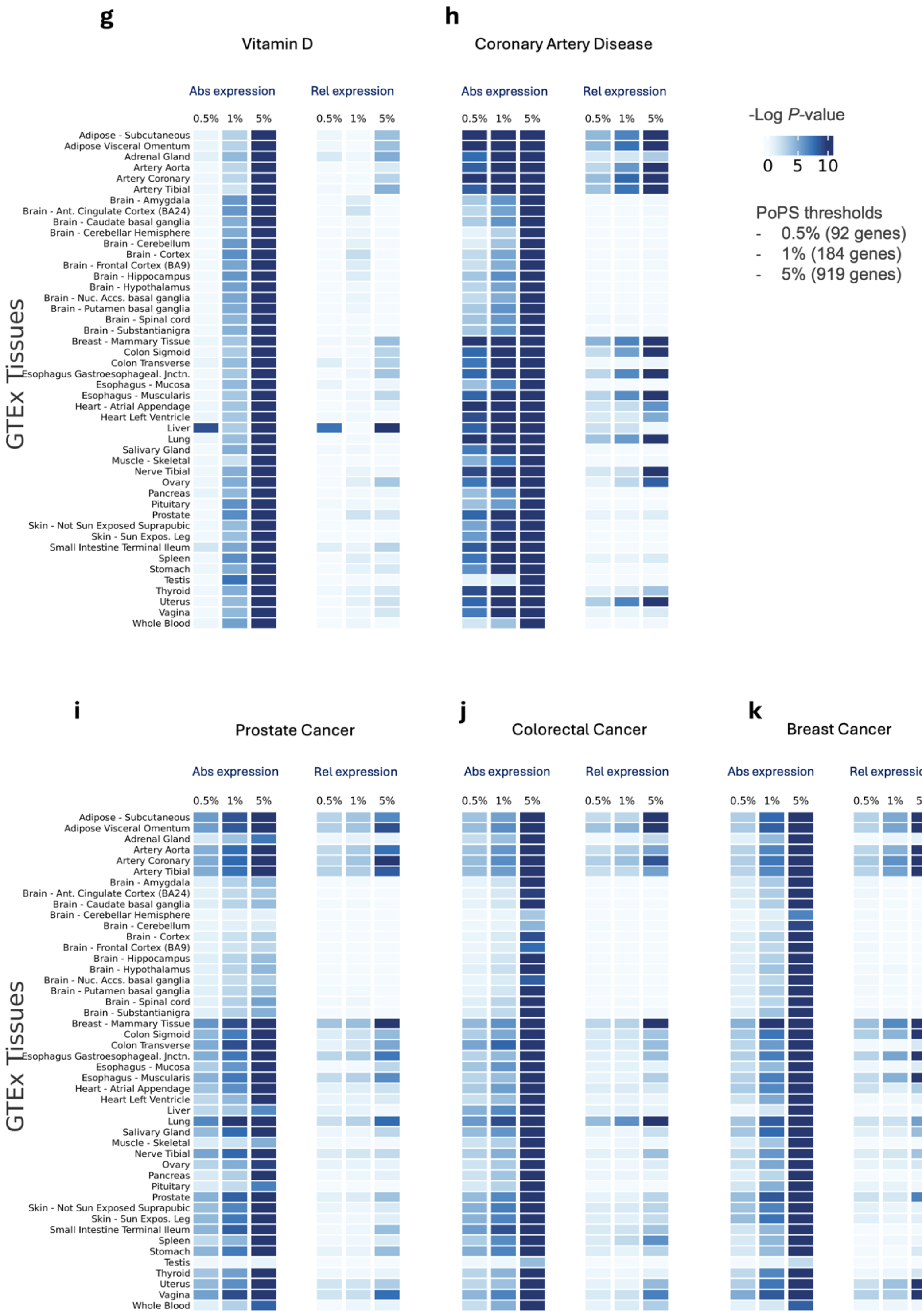


**Supplemental Figure 3:** Heatmap showing results of disease-associated genes, defined using the top 0.5% (92 genes), 1% (184 genes), and 5% (919 genes) of PoPS scores, against all other protein-coding genes (controls) across all GTEx tissues. In blue, results showing the $Log_{10}$ *P*-value for a one-side *t*-tests, testing the null hypothesis that disease-associated genes are not more expressed than other protein-coding genes expressed in that tissue. Abs. Exp, Absolute expression; Rel. Relative expression

#### GWAS to Gene Expression Results Using Open Target Genes as Control Groups

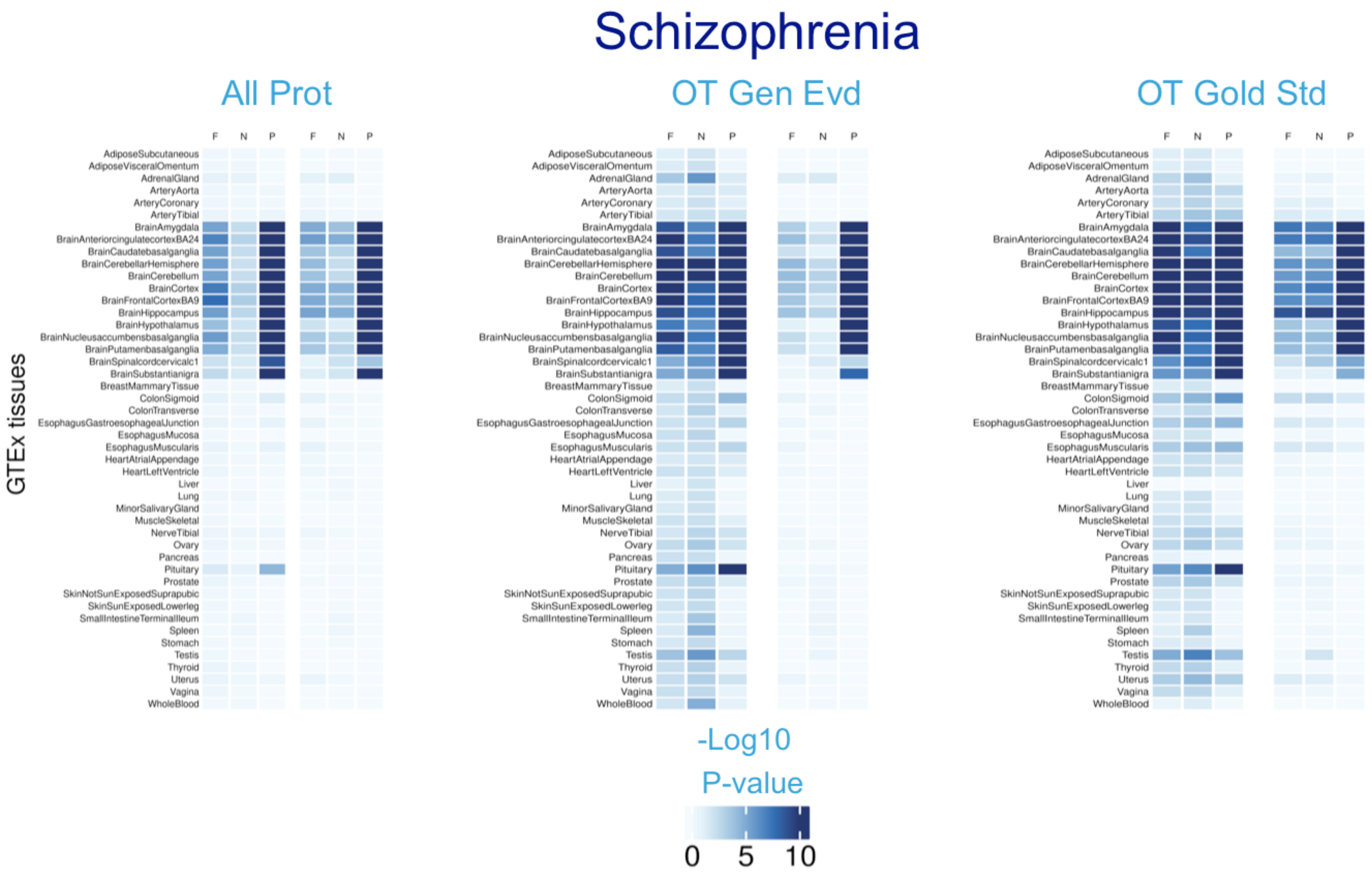


**Supplemental Figure 4**: Heatmaps showing T-test results of the association between gene expression in each GTEx tissue for Schizophrenia. For each disease, three panels are shown showing <u>three different control groups</u> used in the T-test: (i) *"All Prot":* all protein-coding genes, (ii) *"OT Gen Evd":* genes prioritized via Open Targets Genetics evidence and (iii) *"OT Gold Std":* Open Targets gold-standard evidence genes. Within each panel, the three first columns represent results for *Absolute* gene expression, and the second three columns represent *Specific* gene expression. F: Fine-mapped genes, N: Nearest-to-hit genes, P: PoPS genes.

GWAS To Gene Expression Results Using Open Target Genes as Control Groups

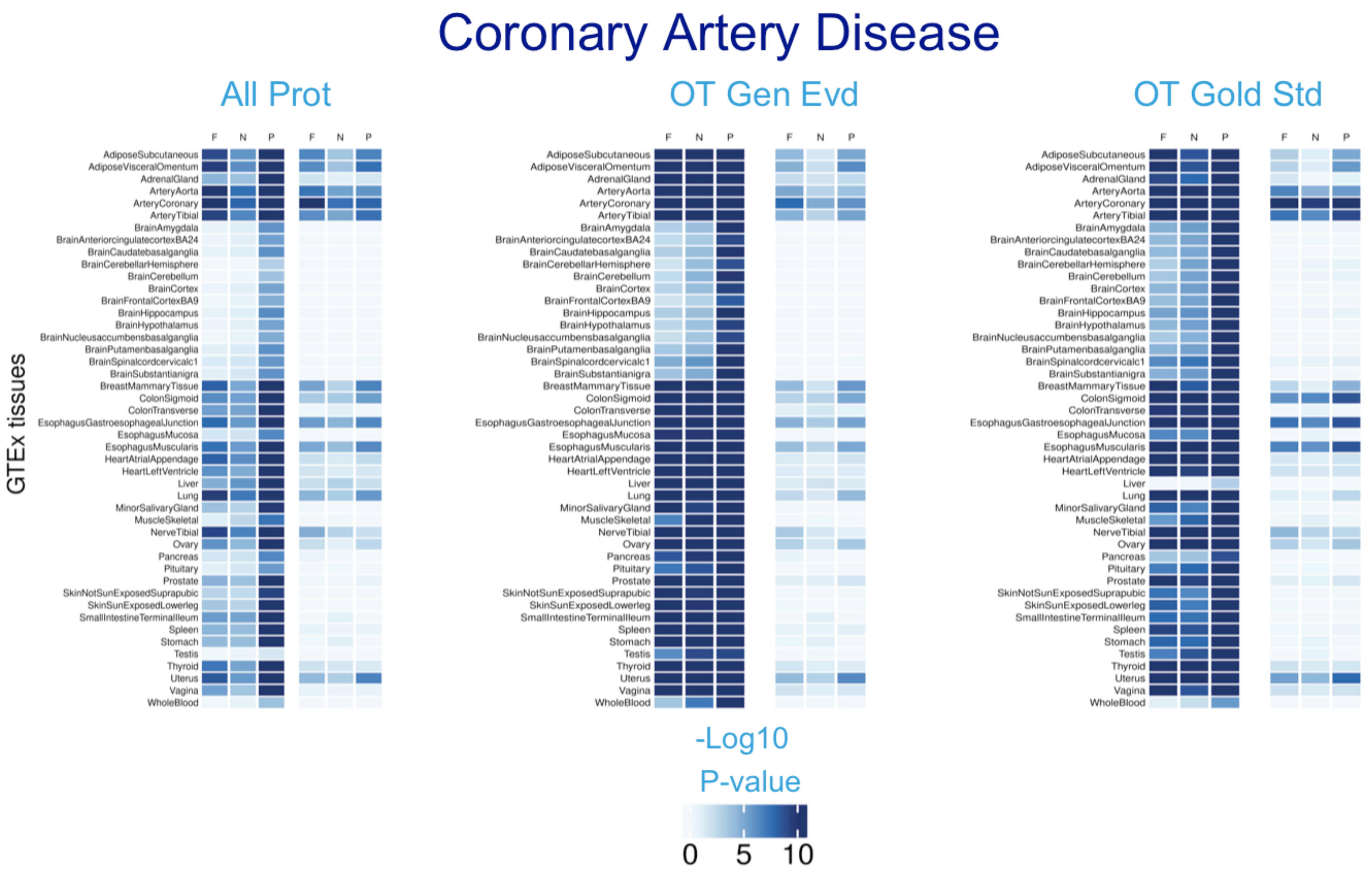


**Supplemental Figure 5**: Heatmaps showing T-test results of the association between gene expression in each GTEx tissue for Coronary Artery Disease. For each disease, three panels are shown showing <u>three different control groups</u> used in the T-test: (i) *"All Prot":* all protein-coding genes, (ii) *"OT Gen Evd":* genes prioritized via Open Targets Genetics evidence and (iii) *"OT Gold Std":* Open Targets gold-standard evidence genes. Within each panel, the three first columns represent results for *Absolute* gene expression, and the second three columns represent *Specific* gene expression. F: Fine-mapped genes, N: Nearest-to-hit genes, P: PoPS genes.

GWAS To Gene Expression Results Using Open Target Genes as Control Groups

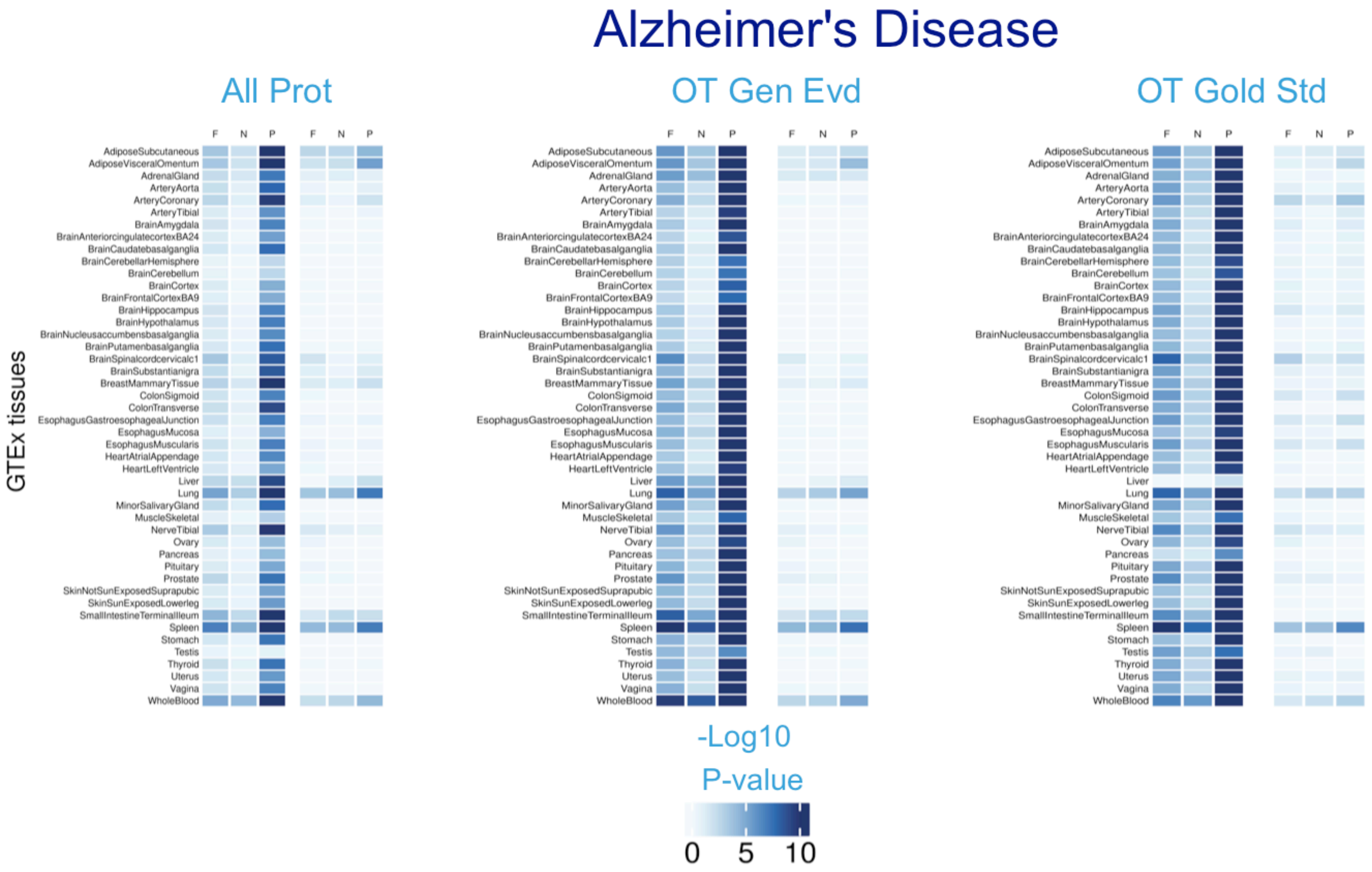


**Supplemental Figure 6**: Heatmaps showing T-test results of the association between gene expression in each GTEx tissue for Alzheimer's Disease. For each disease, three panels are shown showing three different control groups used in the T-test: (i) *"All Prot":* all protein-coding genes, (ii) *"OT Gen Evd":* genes prioritized via Open Targets Genetics evidence and (iii) *"OT Gold Std":* Open Targets gold-standard evidence genes. Within each panel, the three first columns represent results for *Absolute* gene expression, and the second three columns represent *Specific* gene expression. F: Fine-mapped genes, N: Nearest-to-hit genes, P: PoPS genes.

GWAS To Gene Expression Results Using Open Target Genes as Control Groups

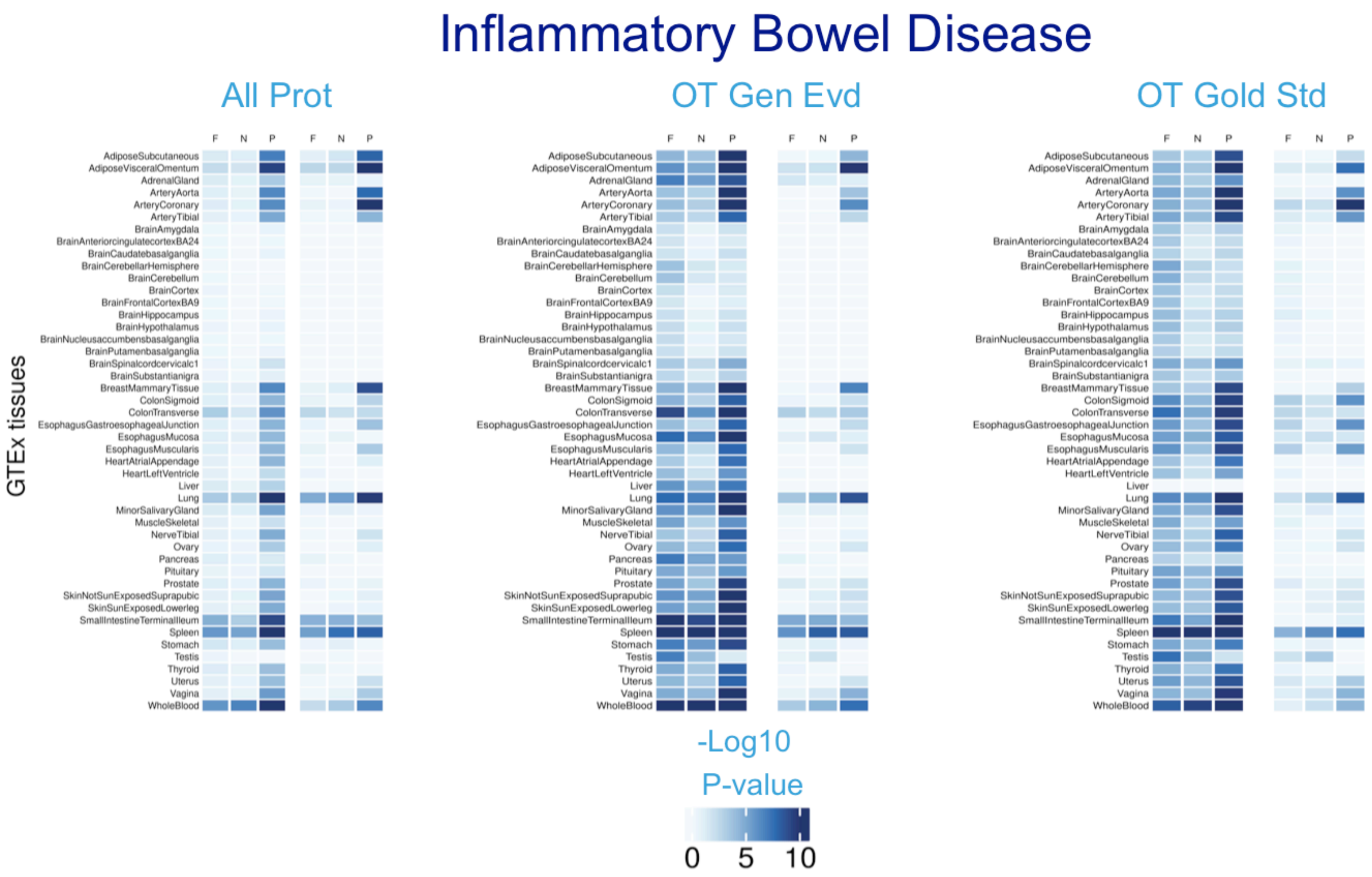


**Supplemental Figure 7**: Heatmaps showing T-test results of the association between gene expression in each GTEx tissue for Inflammatory Bowel Disease. For each disease, three panels are shown showing three different control groups used in the T-test: (i) *"All Prot":* all protein-coding genes, (ii) *"OT Gen Evd":* genes prioritized via Open Targets Genetics evidence and (iii) *"OT Gold Std":* Open Targets gold-standard evidence genes. Within each panel, the three first columns represent results for *Absolute* gene expression, and the second three columns represent *Specific* gene expression. F: Fine-mapped genes, N: Nearest-to-hit genes, P: PoPS genes.

GWAS To Gene Expression Results Using Open Target Genes as Control Groups

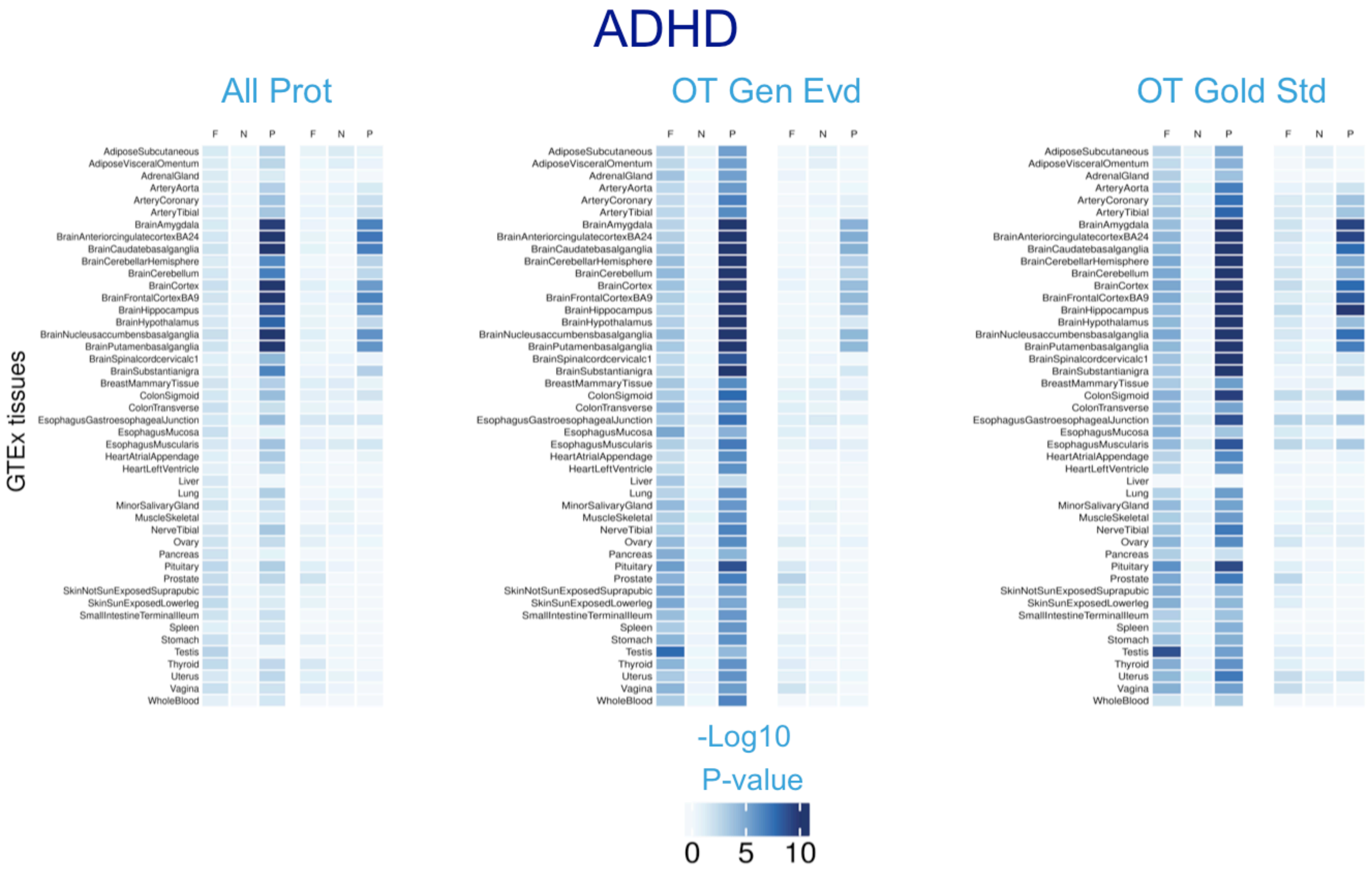


**Supplemental Figure 8**: Heatmaps showing T-test results of the association between gene expression in each GTEx tissue for ADHD. For each disease, three panels are shown showing three different control groups used in the T-test: (i) *"All Prot"*: all protein-coding genes, (ii) *"OT Gen Evd"*: genes prioritized via Open Targets Genetics evidence and (iii) *"OT Gold Std"*: Open Targets gold-standard evidence genes. Within each panel, the three first columns represent results for *Absolute* gene expression, and the second three columns represent *Specific* gene expression. F: Fine-mapped genes, N: Nearest-to-hit genes, P: PoPS genes.

GWAS To Gene Expression Results Using Open Target Genes as Control Groups

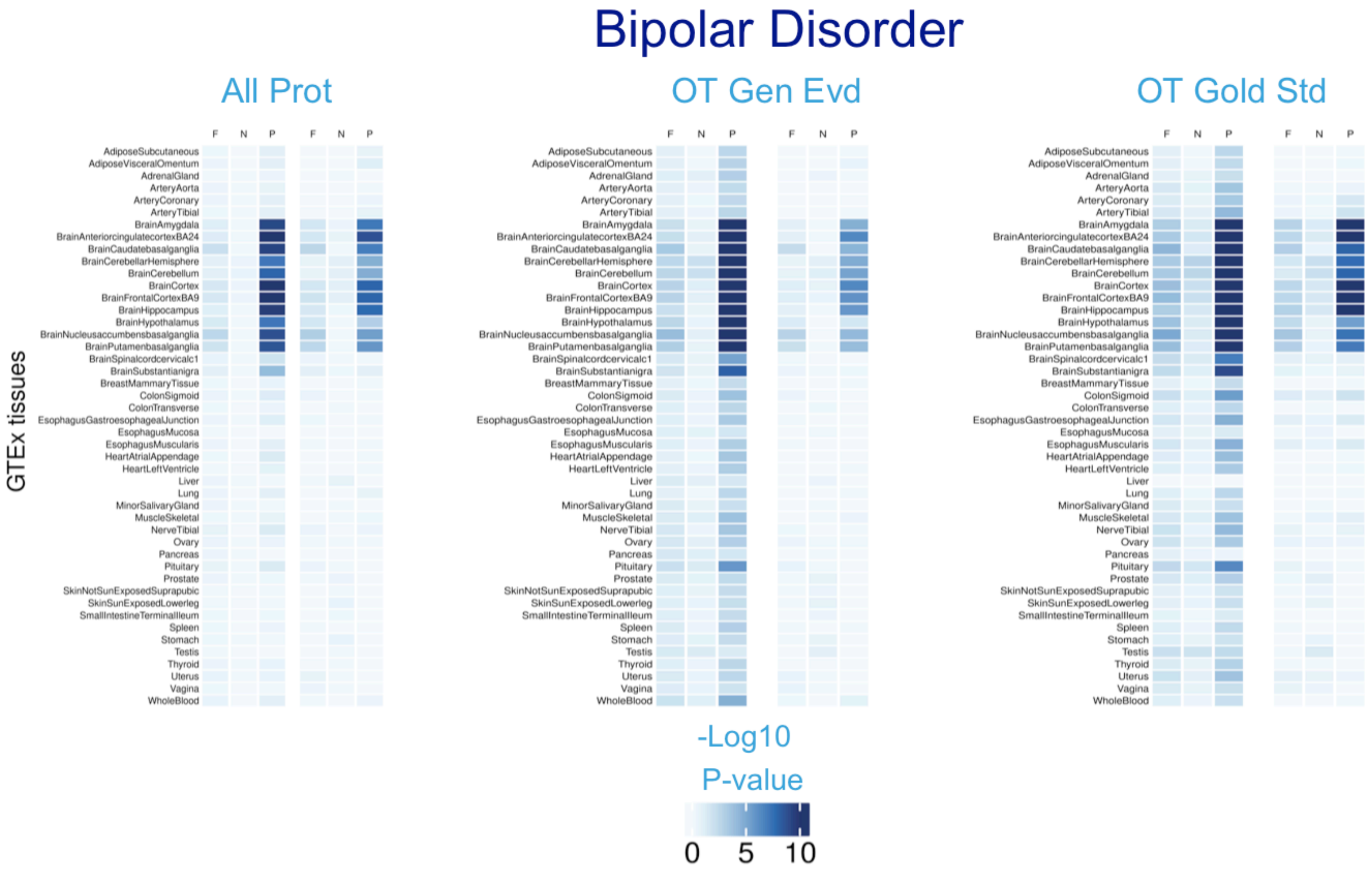


**Supplemental Figure 9**: Heatmaps showing T-test results of the association between gene expression in each GTEx tissue for Bipolar Disorder. For each disease, three panels are shown showing <u>three different control groups</u> used in the T-test: (i) *“All Prot”:* all protein-coding genes, (ii) *“OT Gen Evd”:* genes prioritized via Open Targets Genetics evidence and (iii) *“OT Gold Std”:* Open Targets gold-standard evidence genes. Within each panel, the three first columns represent results for *Absolute* gene expression, and the second three columns represent *Specific* gene expression. F: Fine-mapped genes, N: Nearest-to-hit genes, P: PoPS genes.

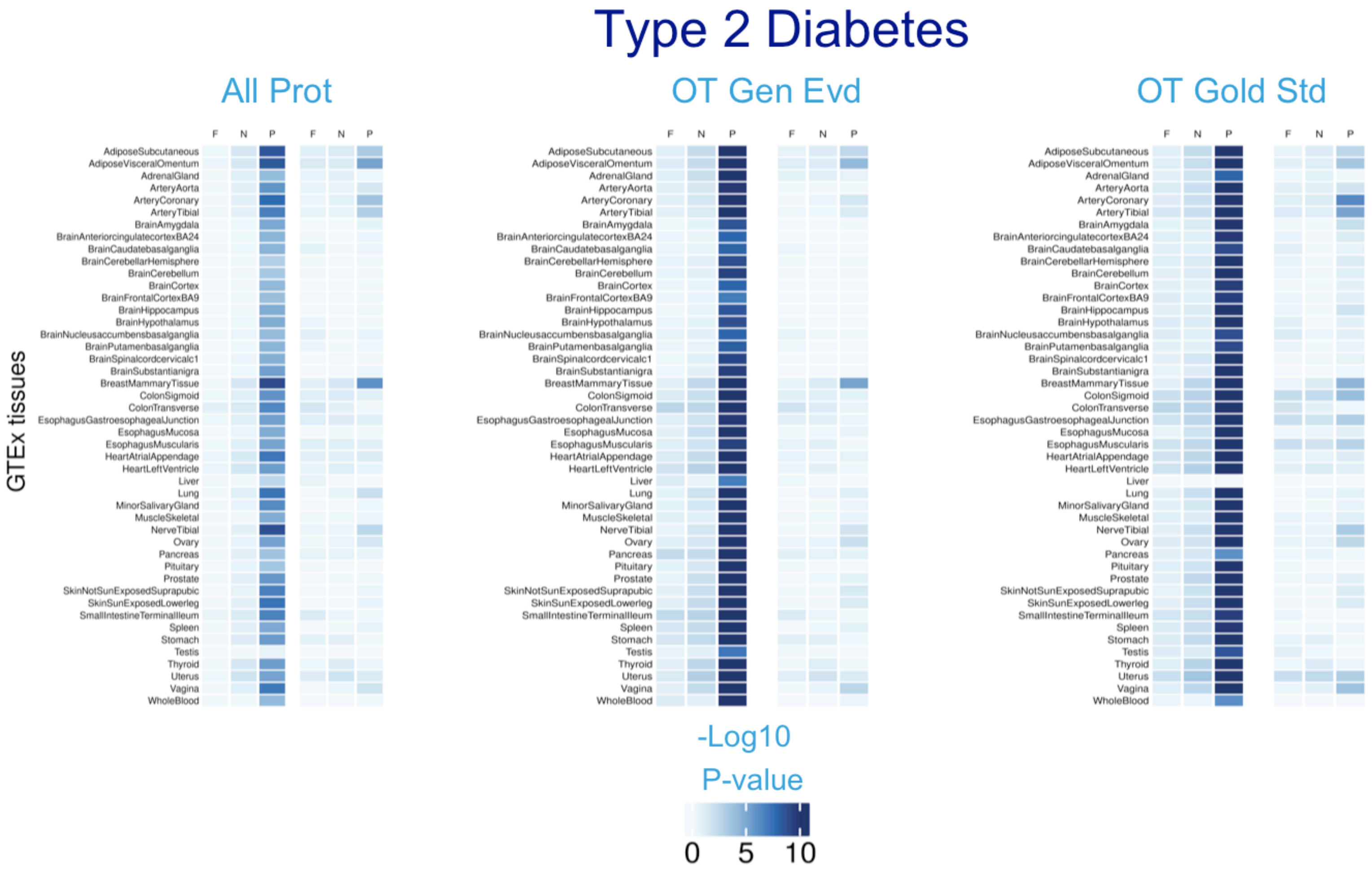


**Supplemental Figure 10**: Heatmaps showing T-test results of the association between gene expression in each GTEx tissue for Type 2 Diabetes. For each disease, three panels are shown showing three different control groups used in the T-test: (i) *"All Prot":* all protein-coding genes, (ii) *"OT Gen Evd":* genes prioritized via Open Targets Genetics evidence and (iii) *"OT Gold Std":* Open Targets gold-standard evidence genes. Within each panel, the three first columns represent results for *Absolute* gene expression, and the second three columns represent *Specific* gene expression. F: Fine-mapped genes, N: Nearest-to-hit genes, P: PoPS genes.

GWAS To Gene Expression Results Using Open Target Genes as Control Groups

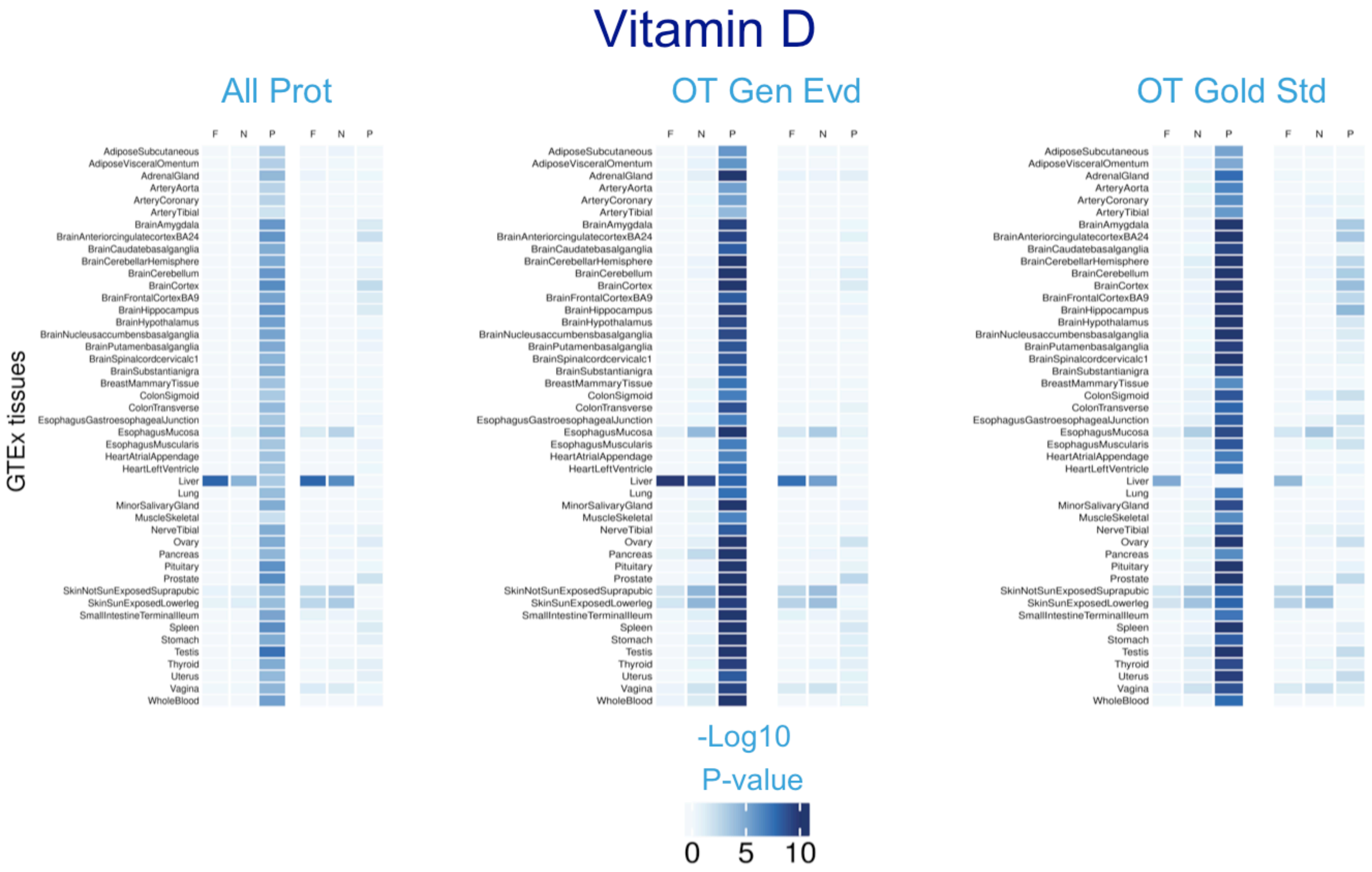


**Supplemental Figure 11**: Heatmaps showing T-test results of the association between gene expression in each GTEx tissue for Vitamin D. For each disease, three panels are shown showing three different control groups used in the T-test: (i) *"All Prot":* all protein-coding genes, (ii) *"OT Gen Evd":* genes prioritized via Open Targets Genetics evidence and (iii) *"OT Gold Std":* Open Targets gold-standard evidence genes. Within each panel, the three first columns represent results for *Absolute* gene expression, and the second three columns represent *Specific* gene expression. F: Fine-mapped genes, N: Nearest-to-hit genes, P: PoPS genes.

#### **Sensitivity analysis where highly expressed genes are removed**

To test whether our association results were driven by only a few genes. We removed the genes that are in the top decile of absolute or relative expression in the relevant tissues, and repeated the '*GWAS to gene expression'* analyses. That is, the *t*-test and Anderson-Darling tests with these new lists of disease-associated genes.

Relevant tissues were defined as those showing significant higher expression across all the tests. Then, we defined a *P*-value threshold for association (threshold = 0.05/45 x 3 x 2, corresponding to 45 tissues, 3 lists of gene prioritization approaches, and 2 test statistics (*t*-test and Anderson-Darling test). Then, we identified the tissues for which the Anderson-Darling and the *t*-test showed *P*-values < threshold.

Results show that, while the *P*-values increase for all the tests, results remain consistent after removing the top 10% expressed genes.

### Sensitivity analyses – Removal of highly expressed genes

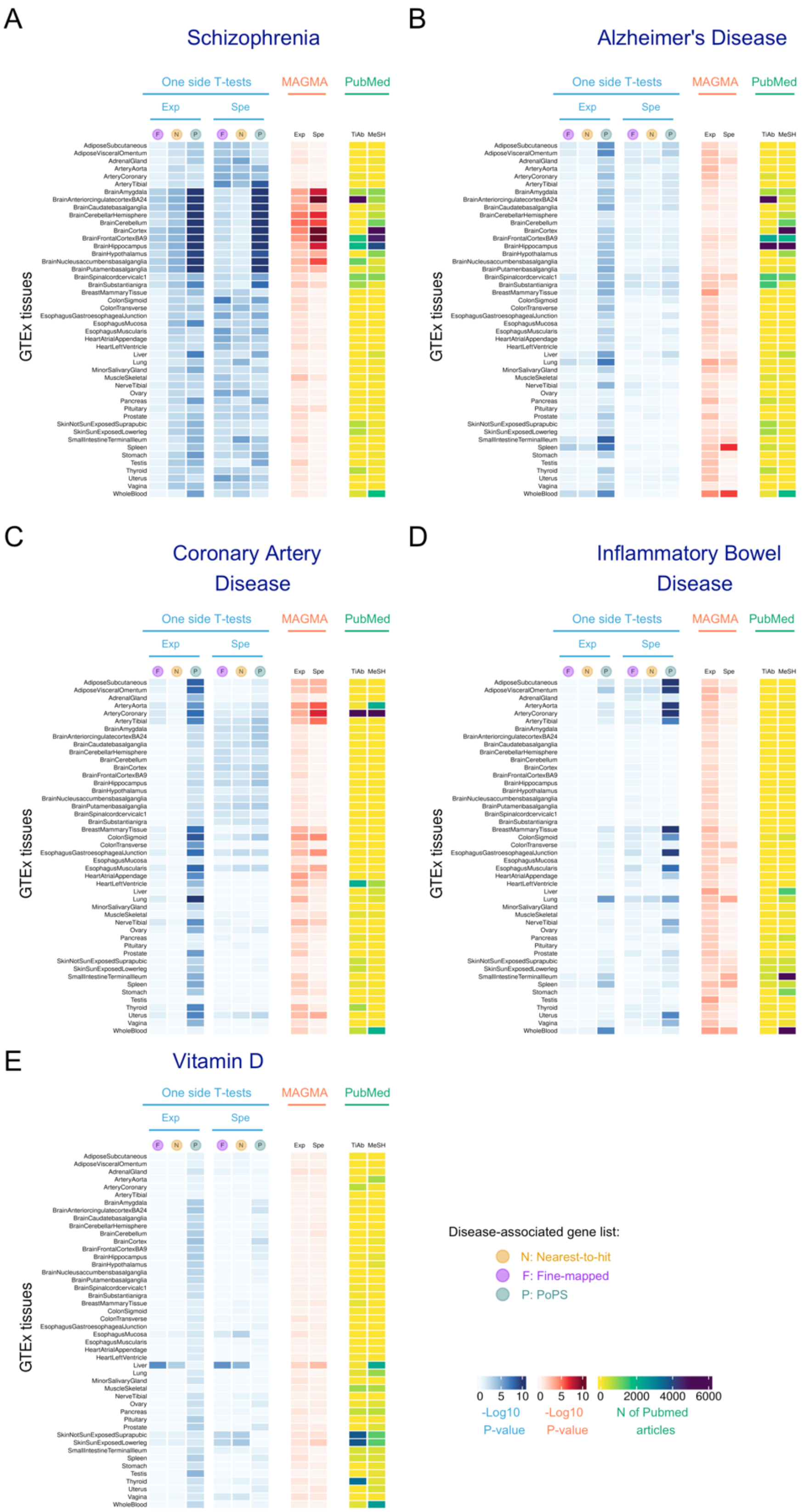


**Supplemental Figure 12.** Heatmap showing results of the sensitivity analysis where association between gene expression in each GTEx tissue was performed after removal of the top 10% genes most expressed or most specifically expressed in relevant tissues.

### The Gene Expression Landscape of Disease Genes in the ARCHS4 database

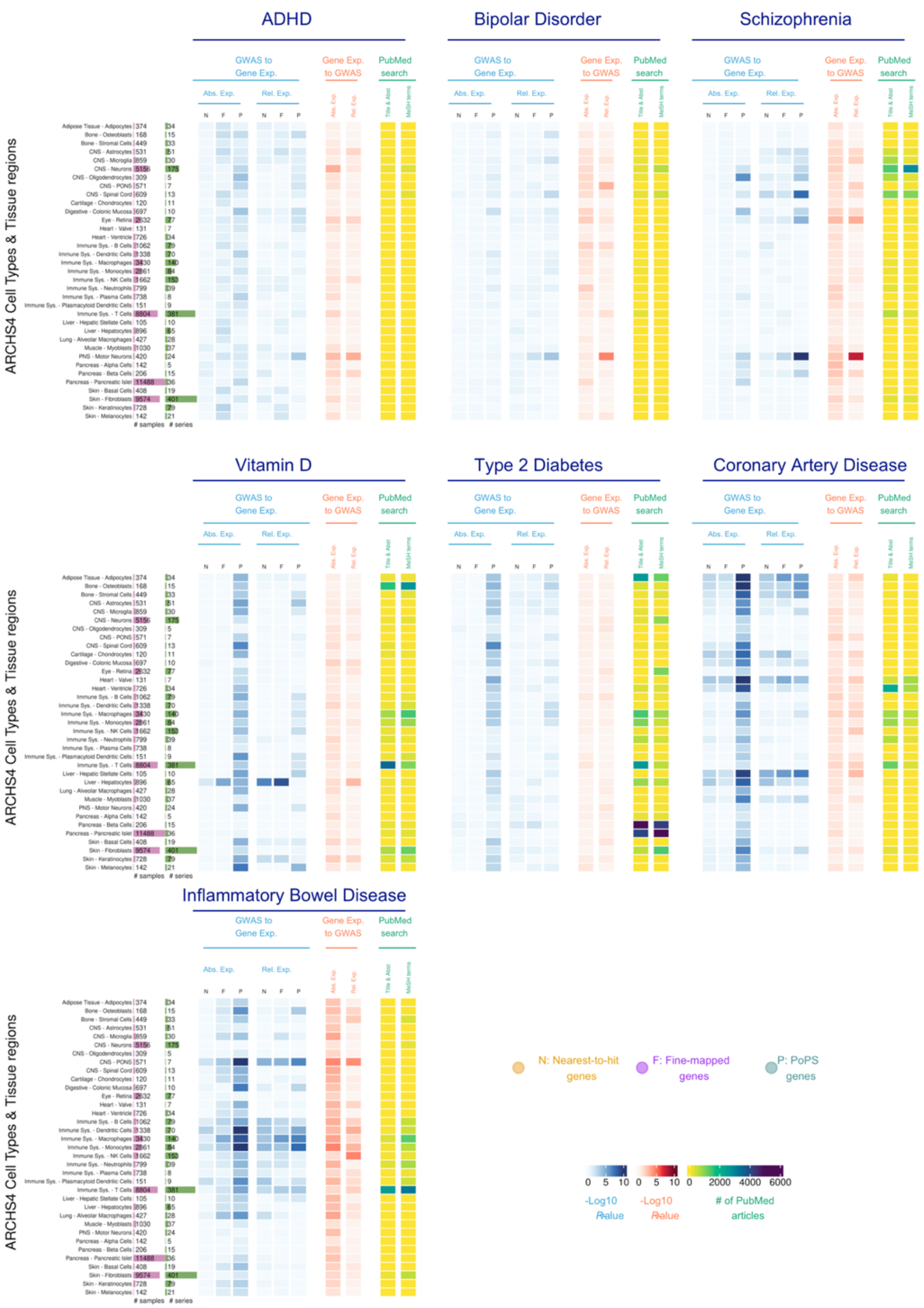


**Supplemental Figure 13**: Heatmap showing results of the association between gene expression in each cell-type or tissue region and ADHD, Bipolar Disorder, Schizophrenia, Vitamin D, and Type 2 Diabetes **using the ARCHS4 dataset**. In blue, results showing the $Log_{10}$ *P*-value for a one-side T-tests. In red, results showing the $Log_{10}$ *P*-value for enrichment of GWAS signal across the set of genes with highest absolute and relative expression for each tissue. In yellow, results for the Literature Search using PubMed. Abs. Exp, Absolute expression; Spec. Exp, specific expression; F, Fine-mapped genes; N, Nearest-to-hit genes, P, Polygenic Priority Scores genes; Title & Abst, Title and Abstract are used in the PubMed Search; MeSH, MeSH terms are used in the PubMed Search.

#### Analysis of eight common cancer traits

*Cancer traits with all prioritized gene lists available*

For three cancer traits (breast, prostate, and colorectal cancer) fine-mapped, PoPS and GWAS data were available. Therefore, we applied the three main approaches (1) GWAS → Gene Expression, (2) Gene Expression → GWAS, and (3) PubMed Search, in all the gene expression resources used in this study: GTEx (all tissues), ARCHS4 (all cell types), and Tabula Sapiens (prostate, mammary, small, and large intestine). Gene lists for each prioritization method used for these three cancer types are provided in **Additional File 2, Supplemental Tables 1-3**. Prioritized genes were defined as follows:

- *Definition of 'nearest-to-hit' cancer genes*: To identify genes closest to GWAS hits, we used publicly available GWAS summary statistics for the three cancers studied. Processing was performed as for the major diseases studied. Clumping was performed in PLINK 1.9 with 1000 Genomes data as the LD reference. Variants with $P \leq 5\times10^{-8}$ were retained, while those within 250 Kbp and $r^2 \geq 0.5$ or with $P \geq 0.01$ were removed. The nearest protein-coding gene to each index variant was identified using ENSEMBL GRCh37.75 gene coordinates. The table below provides details on GWAS sources, clumped variants, identified genes, and variant-gene distances.

**Table**. Gene prioritization based on the nearest gene to GWAS hit. Table shows references for each GWAS summary statistics, the number of clumped SNPs, genes and median distance between SNP and selected gene.

| Cancer type | GWAS summary statistics used | GWAS catalogue accession | N clumped SNPs | N unique ensemble IDs | Median distance between SNP and nearest protein coding gene (bp) |
|---|---|---|---|---|---|
| Breast | Zhang et al 2020[12] | GCST90454347 | 103 | 29 | 27,444 |
| Prostate | Wang et al 2023[13] | GCST90274713 | 3974 | 852 | 19,310 |
| Colorectal | Fernandez-Rozadilla et al 2022[14] | GCST90129505 | 282 | 235 | 37,875 |

The gene expression landscape of cancer genes

- *Definition of 'fine-mapped' cancer genes*: Gene lists previously fine-mapped and prioritized using GWAS for Breast[15], Prostate[16] and Colorectal[17] cancer were obtained. These gene lists were created integrating statistical fine-mapping with functional datasets on the cancer disease tissue of interest. The number of genes prioritized were 179 for Breast cancer, 447 for prostate cancer, and 177 for Colorectal cancer.

- *Definition of 'PoPS cancer genes'*: Similar to analyses for major diseases, we extracted genes with the top 1% PoPS scores, resulting in a list of 184 prioritized genes per cancer type. Results using top 0.5% and 5% PoPS scores yielded consistent results (**Supplemental Figure 3**).

Venn diagrams in **Supplemental Figures 14-16** illustrate the overlap of genes among the three prioritization approaches. Consistent with findings in major common diseases, the overlap was low: only four genes for breast cancer, six for prostate cancer, and two for colorectal cancer were prioritized by all three methods. Data processing and statistical analyses followed the same pipeline used for major diseases (detailed in the main text).

GTEx analyses revealed strong associations between cancer-associated genes and the expected relevant tissues: Results for *t*-tests (blue columns in **Supplemental Figures 14-16**), show that genes linked to prostate cancer were highly expressed in the prostate ($P$-values < $10^{-9}$), and fine-mapped and PoPS genes associated with breast and colorectal cancer were more expressed in mammary tissue ($P$-values < $10^{-10}$), and transverse colon ($P$-values < $10^{-8}$), respectively. Similar to results for eight major diseases in the main text, PoPS-prioritized genes showed smaller $P$-values across a broader range of tissues, particularly when absolute gene expression was considered. In contrast, for breast and colorectal cancer, the nearest-gene approach did not yield significant associations with expected tissues, a pattern also observed in the Gene Expression to GWAS method (red columns in **Supplemental Figures 14-16**). Note that the number of genes for these two cancers (29 for breast cancer, and 235 for colorectal cancer) is lower compared to the number of Nearest genes for Prostate cancer (852 genes).

Beyond expected tissue associations, cancer-associated genes were also linked to other tissues (**Additional File 3, Supplemental Table 1**). For example, fine-mapped genes for breast cancer showed increased relative expression across skin (*P*-values = $7.35 \times 10^{-14}$), nerve tibial (*P*-values = $2.36 \times 10^{-15}$), arteries (*P*-values < $10^{-13}$) and thyroid (*P*-values 1.08 < $10^{-13}$), as well as higher absolute expression for multiple brain tissues (*P*-values < $10^{-12}$). Prostate cancer-associated genes were also more highly expressed in the vagina, uterus, and salivary gland (*P*-values < $10^{-6}$ across approaches). Genes associated with colorectal cancer showed increased expression across a range of non-brain tissues, but these were only observed for fine-mapped and PoPS genes. Although the ARCHS4 dataset does not include direct cancer-affected tissues, it provided additional resolution for these unexpected findings (**Additional File 3, Supplemental Table 2**). For example, fine-mapped breast and colorectal cancer genes were significantly more expressed in GTEx skin, a pattern also observed in ARCHS4, where fine-mapped and PoPS genes showed higher expression in basal cells and keratinocytes (*P*-values < $10^{-6}$). Similarly, PoPS-prioritized colorectal cancer genes exhibited higher specificity in the liver in GTEx and were also highly expressed in hepatocytes and hepatic stellate cells in ARCHS4 (*P*-values < $10^{-4}$).

For the Tabula Sapiens analyses, we examined gene expression in mammary, prostate, and intestinal tissues using the same single-cell RNA-seq processing pipeline as for major diseases to provide further resolution of the cancer tissue-specific associations (**Additional File 3, Supplemental Table 3**).

Overall concordance in results across gene prioritization methods was low, being higher between the nearest and fine-mapped genes for prostate cancer, where epithelial subtypes - including luminal, CPE-high luminal, and CGB3A1-high club cells- showed higher expression of prostate cancer genes (*P*-values < $10^{-4}$).

Breast cancer results varied significantly by gene prioritization method, but across fine-mapped and nearest-gene prioritization methods, as well as the Gene Expression to GWAS approach (**Additional File 3, Supplemental Table 4**), B cells and mature luminal cells consistently showed significant associations in relative expression (*P*-values < $10^{-3}$).

For colorectal cancer, none of the immune cell types included in ARCHS4 showed increased expression of immune cell types. However, in the Tabula Sapiens dataset, fibroblasts, monocytes, and neutrophils had the lowest P-values (*P*-values < $10^{-4}$), suggesting the potential relevance of immune cell types in disease mechanisms. This is in line with the PubMed Literature results, which suggests immune cell types have been broadly studied for this cancer.

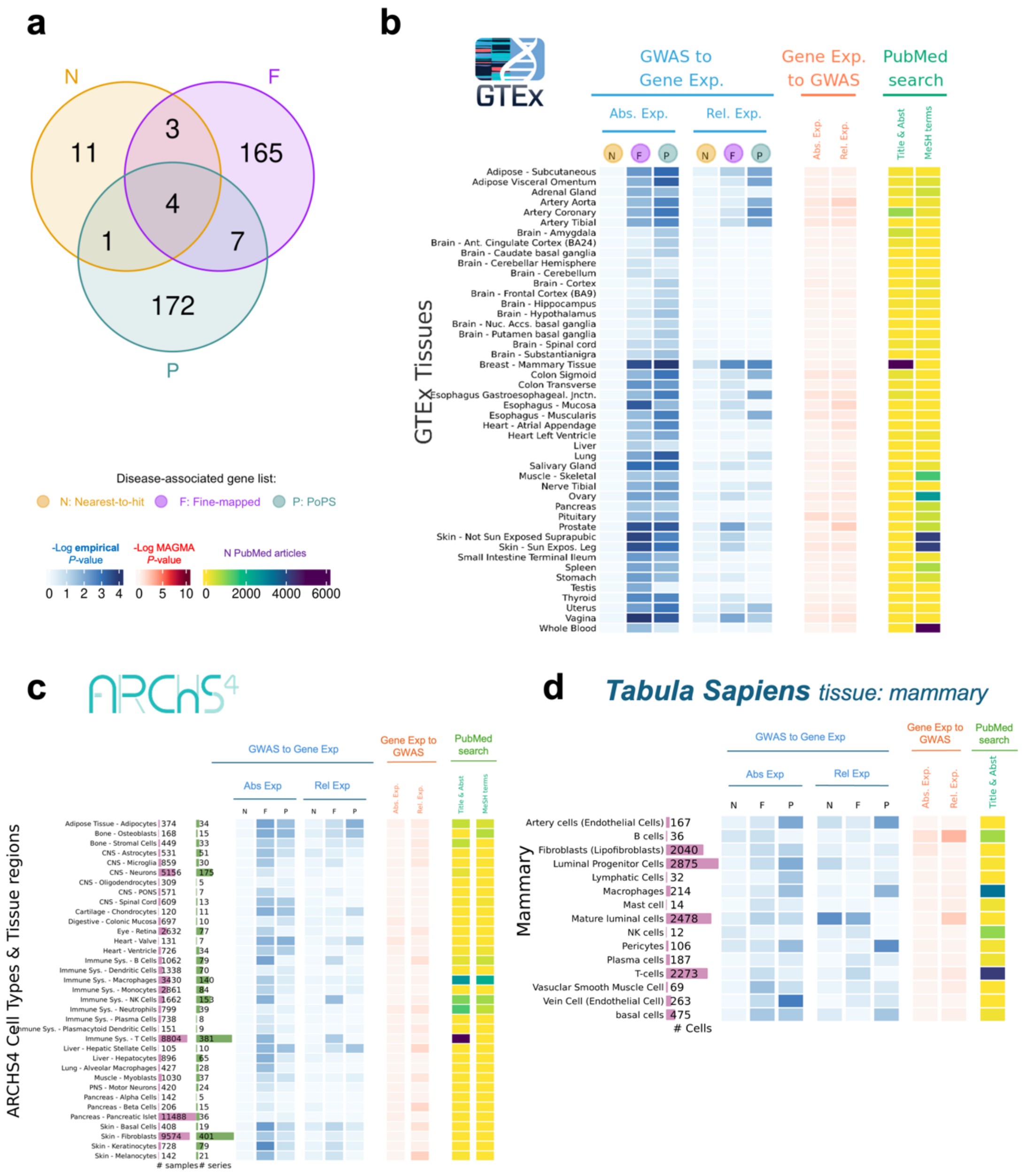

**Supplemental Figure 14**. The Gene expression landscape of genes associated with Breast Cancer. **a**, Venn diagrams showing the overlap of genes for each of the gene prioritization approaches included; **b-d)** Heatmap showing results of the association between breast cancer genes and gene expression in: **b**, each GTEx tissue; **c**, ARCHS4 cell types; **d**, Tabula Sapiens cell types in mammary tissue. In blue, results showing the $\text{Log}_{10}$ *P*-value for a one-side *t*-tests (for ARCHS4 and GTEx) and empirical *P*-value for the Tabula Sapiens dataset (10000 permutations), testing the null hypothesis that disease-associated genes are not more expressed than other protein-coding genes expressed in that tissue. In red, results showing the $\text{Log}_{10}$ *P*-value for enrichment of GWAS signal across the set of genes with highest absolute and relative expression for each tissue. In yellow, results for the Literature Search using PubMed. Abs. Exp, Absolute expression; Rel. Relative expression; F, Fine-mapped genes; N, Nearest-to-hit genes, P, Polygenic Priority Scores genes; Title & Abst, Title and Abstract; MeSH, MeSH terms.

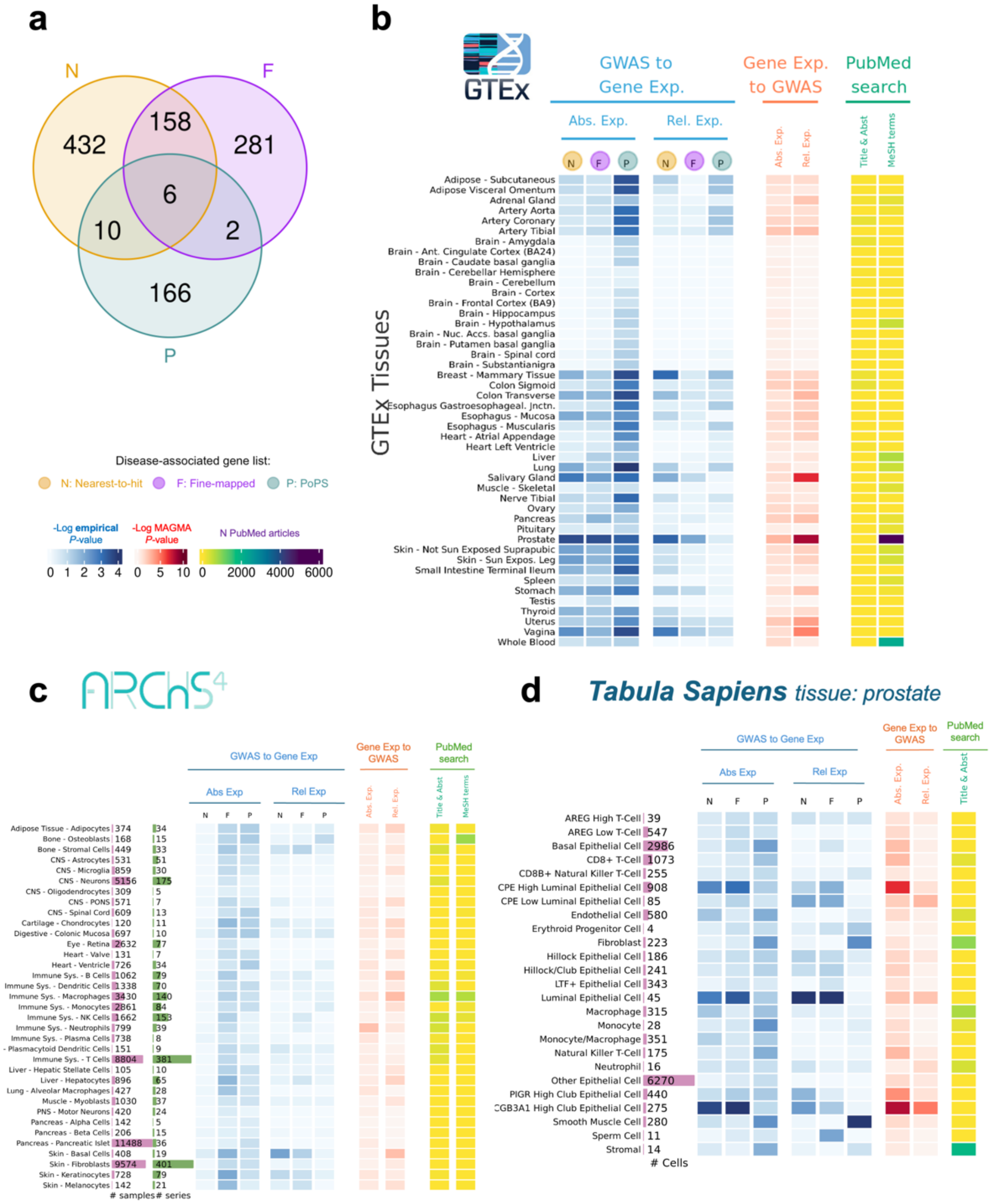


**Supplemental Figure 15**. The Gene expression landscape of genes associated with Prostate Cancer. **a**, Venn diagrams showing the overlap of genes for each of the gene prioritization approaches included; **b-d)** Heatmap showing results of the association between breast cancer genes and gene expression in: **b**, each GTEx tissue; **c**, ARCHS4 cell types; **d**, Tabula Sapiens cell types in prostate tissue. In blue, results showing the $\text{Log}_{10}$ *P*-value for a one-side *t*-tests (for ARCHS4 and GTEx) and empirical *P*-value for the Tabula Sapiens dataset (10000 permutations). In red, results showing the $\text{Log}_{10}$ *P*-value for enrichment of GWAS signal across the set of genes with highest absolute and relative expression for each tissue. In yellow, results for the Literature Search using PubMed. Abs. Exp, Absolute expression; Rel. Relative expression; F, Fine-mapped genes; N, Nearest-to-hit genes, P, Polygenic Priority Scores genes; Title & Abst, Title and Abstract; MeSH, MeSH terms.

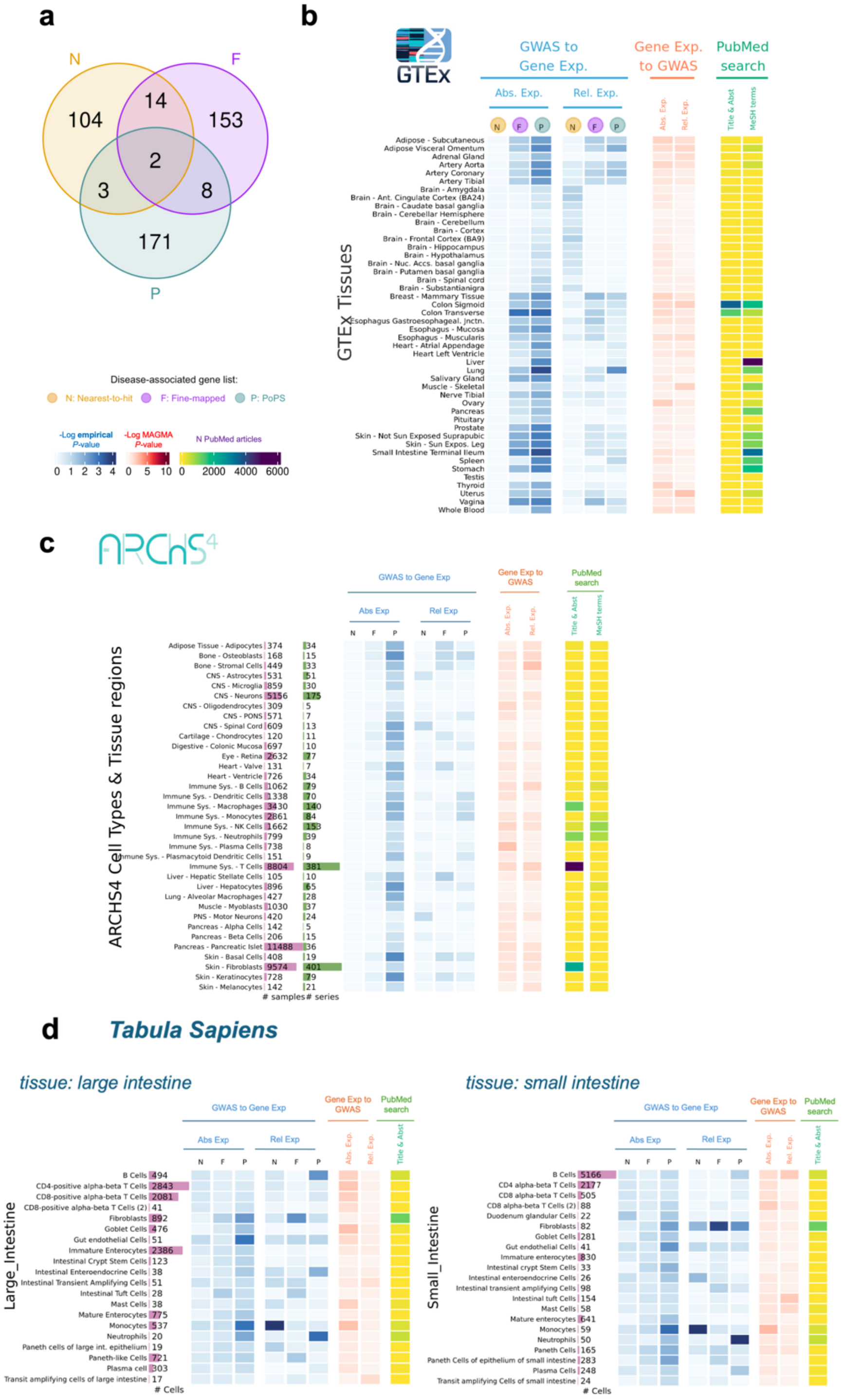


**Supplemental Figure 16**. The Gene expression landscape of genes associated with Colorectal Cancer. **a**, Venn diagrams showing the overlap of genes for each of the gene prioritization approaches included; **b-d)** Heatmap showing results of the association between breast cancer genes and gene expression in: **b**, each GTEx tissue; **c**, ARCHS4 cell types; **d**, Tabula Sapiens cell types in small and large intestine tissue. In blue, results showing the $\text{Log}_{10}$ *P*-value for a one-side *t*-tests (for ARCHS4 and GTEx) and empirical *P*-value for the Tabula Sapiens dataset (10000 permutations). In red, results showing the $\text{Log}_{10}$ *P*-value for enrichment of GWAS signal across the set of genes with highest absolute and relative expression for each tissue. In yellow, results for the Literature Search using PubMed. Abs. Exp, Absolute expression; Rel. Relative expression; F, Fine-mapped genes; N, Nearest-to-hit genes, P, Polygenic Priority Scores genes; Title & Abst, Title and Abstract; MeSH, MeSH terms.

#### *Sex stratified analyses for cancer traits affecting sex-specific tissues*

We also performed sex stratified analyses for cancers where the cancerous tissue is only available in one of the sexes (i.e. ovary, prostate, breast), and assessed the expression of disease genes across all GTEx tissues in men and women separately. Results were as expected (**Supplemental Figure 17** and **Additional File 3, Supplemental Table 5**): Despite the low number of genes identified in ovary cancer, the Anderson-Darling test shows a significant association for vagina in women (*P*-value = $7.62 \times 10^{-6}$), and genes related to prostate cancer are highly expressed in men. In addition, the association of breast and breast cancer is less significant in men (Rel. Expression *P*-value=0.0018) than in women (Rel. Expression *P*-value=0.00012).

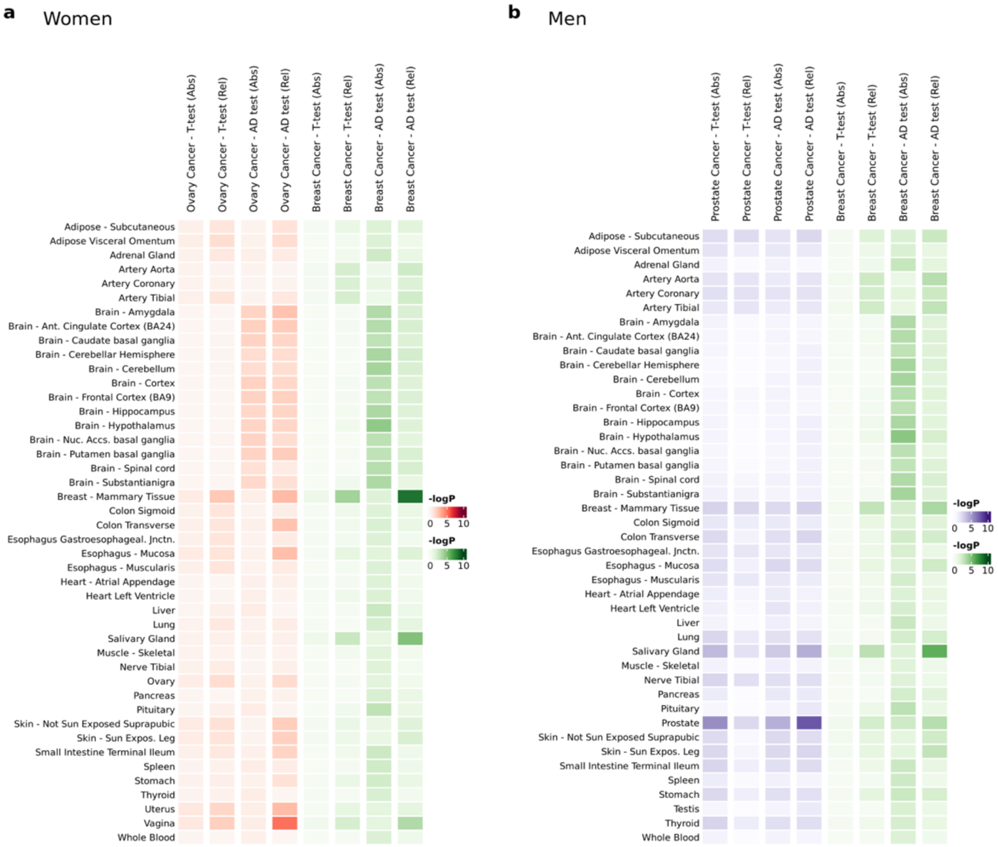


**Supplemental Figure 17**: Sex-stratified analyses for three cancer traits: ovary cancer, breast cancer, and prostate cancer. Heatmaps show *P*-values for the T-tests and Anderson-Darling (AD test) results of the association between absolute (Abs) and Relative (Rel) gene expression in each GTEx tissue.

*Cancer traits using only the "nearest-to-hit genes" approach*

For five other cancer traits, we only applied the "nearest-to-hit genes" approach, as fine-mapping and PoPS-based gene lists were unavailable. Here we used a publicly available dataset that included lists of independent, GWAS significant SNPs used to construct polygenic risk scores[18]. The number of independent SNPs were: 19 for lung cancer, 15 for kidney cancer, 14 for bladder cancer, 31 for ovary cancer and 22 for pancreatic cancer (**Additional File 3, Supplemental Table 6**). In contrast to the other diseases analysed in this study, for which we had full summary statistics, these lists of SNPs were already independent. Therefore, we did not perform clumping on them. The procedure for assigning the closest gene to each GWAS hit was the same as for *'nearest-to-hit'* genes.

Results for these five cancer types show non-significant tissue-trait association results (**Supplemental Figure 18**), with all t-test P-values being greater than $10x^{-2}$ (**Additional File 3, Supplemental Table 7**). The strongest association observed was the expression of genes associated with Ovary cancer being more specifically expressed in the vagina. However, this result was only significant when tested with the AD test (*P*-value = $8.80x10^{-6}$).

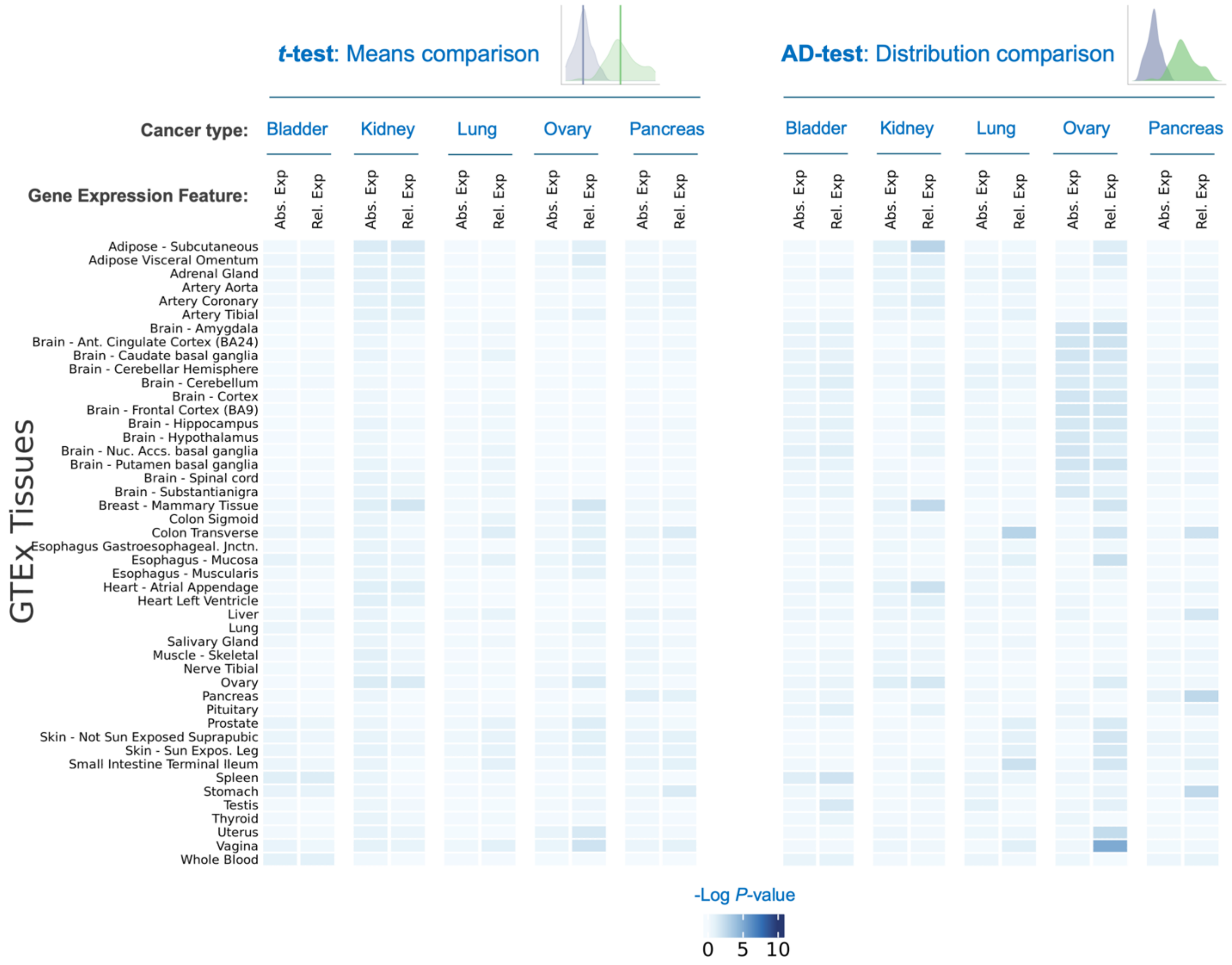

**Supplemental Figure 18**: Heatmaps showing t-tests and Anderson-Darling (AD-test) results of the association between gene expression in each GTEx tissue. Abs. exp, Absolute gene expression was measured; Rel. spe, Specific (relative) gene expression was measured; AD, Anderson-Darling test.

Biological and Technical Factors Influencing Disease Gene Expression (Variance Partition)

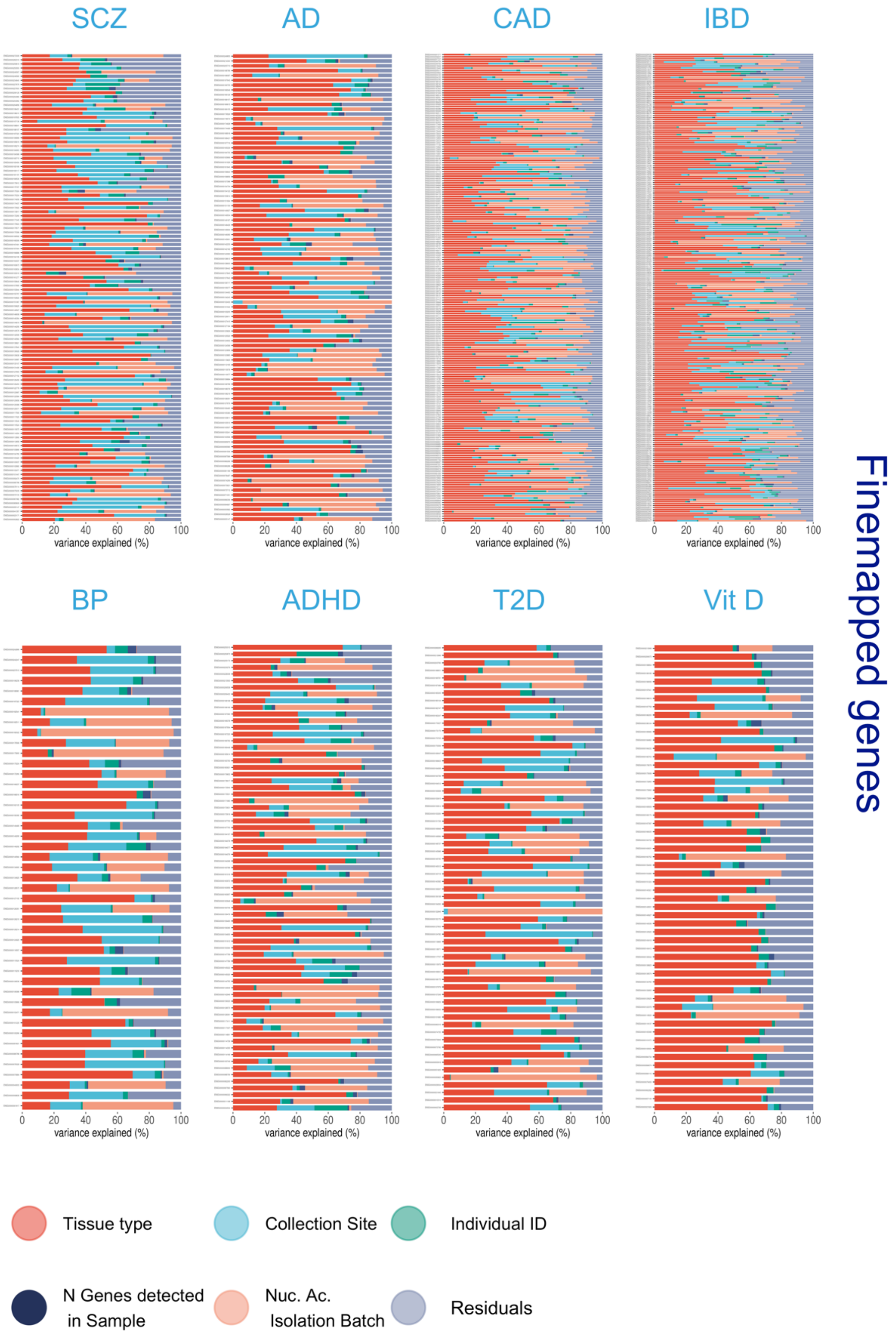


**Supplemental Figure 19**. Factors explaining gene expression variance in disease-associated genes prioritized via fine-mapping of GWAS hits. Each percentage bar (each row) corresponds to an individual gene. Panels are split by prioritization method and trait investigated.

Biological and Technical Factors Influencing Disease Gene Expression (Variance Partition)

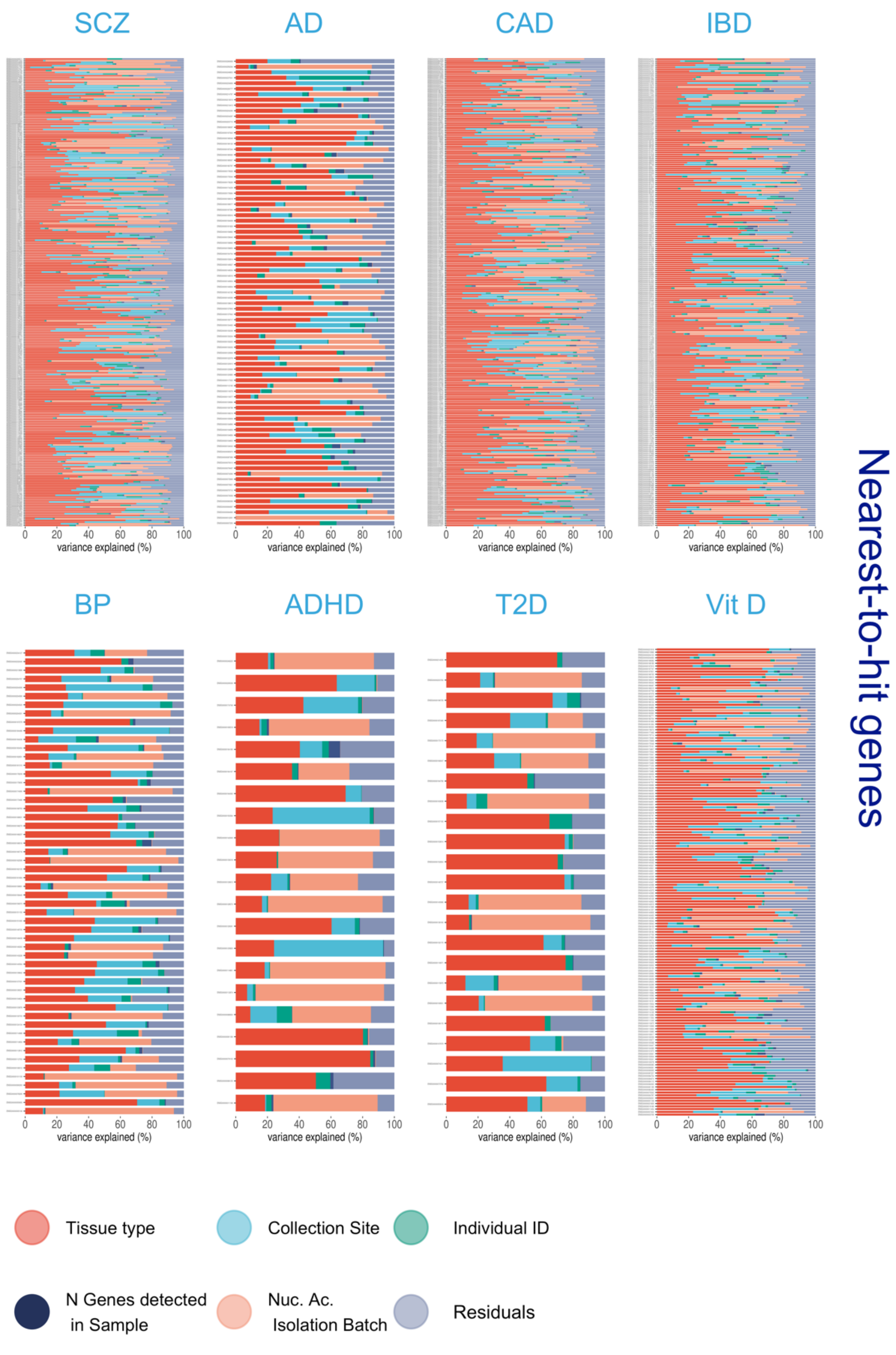


**Supplemental Figure 20**. Factors explaining gene expression variance in disease-associated genes prioritized using the nearest to GWAS-hit approach. Each percentage bar (each row) corresponds to an individual gene. Panels are split by prioritization method and trait investigated.

Biological and Technical Factors Influencing Disease Gene Expression (Variance Partition)

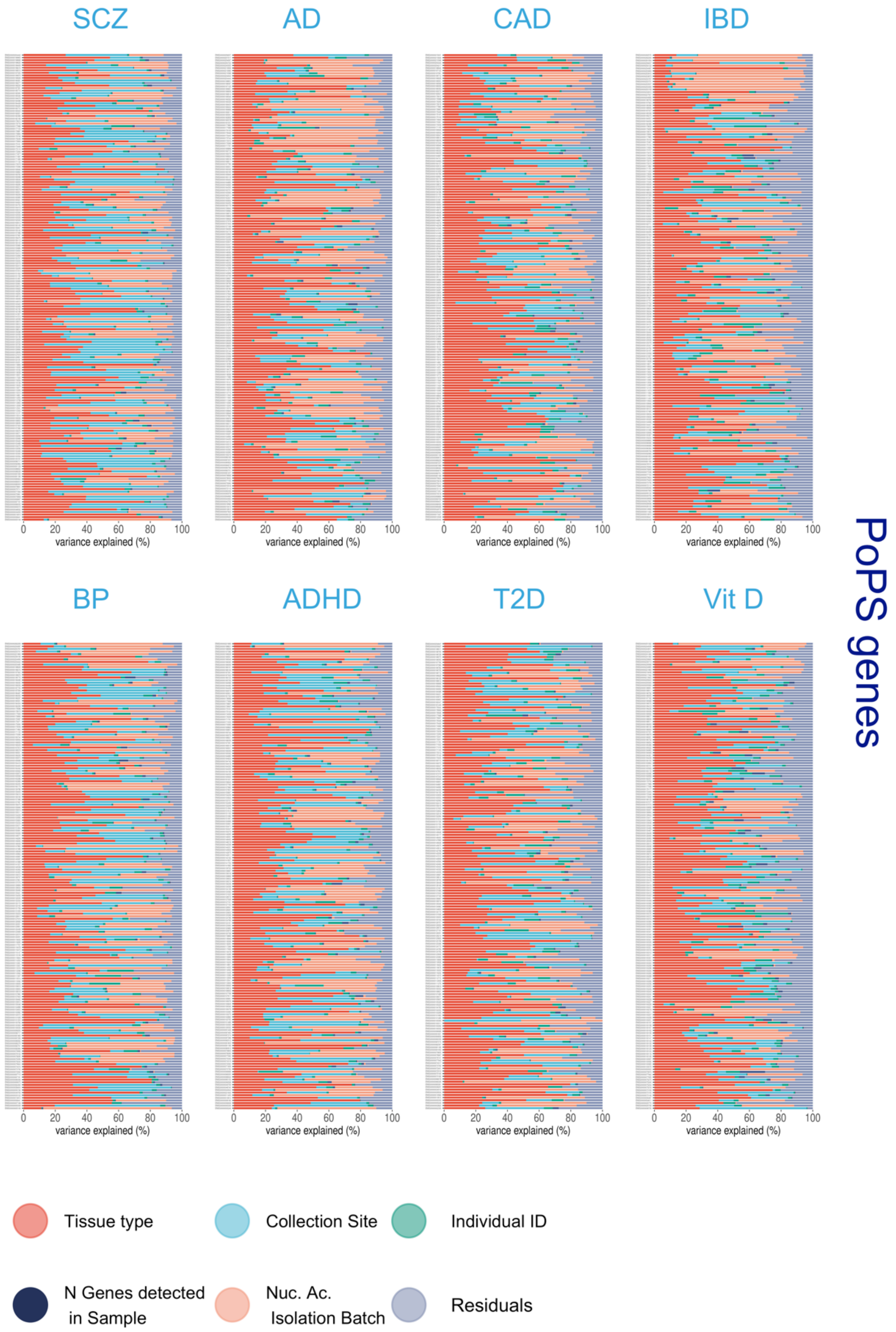


**Supplemental Figure 21**. Factors explaining gene expression variance in disease-associated genes using the PoPS method. Each percentage bar (each row) corresponds to an individual gene. Panels are split by prioritization method and trait investigated.

### Biological and Technical Factors Influencing Disease Gene Expression (Variance Partition)

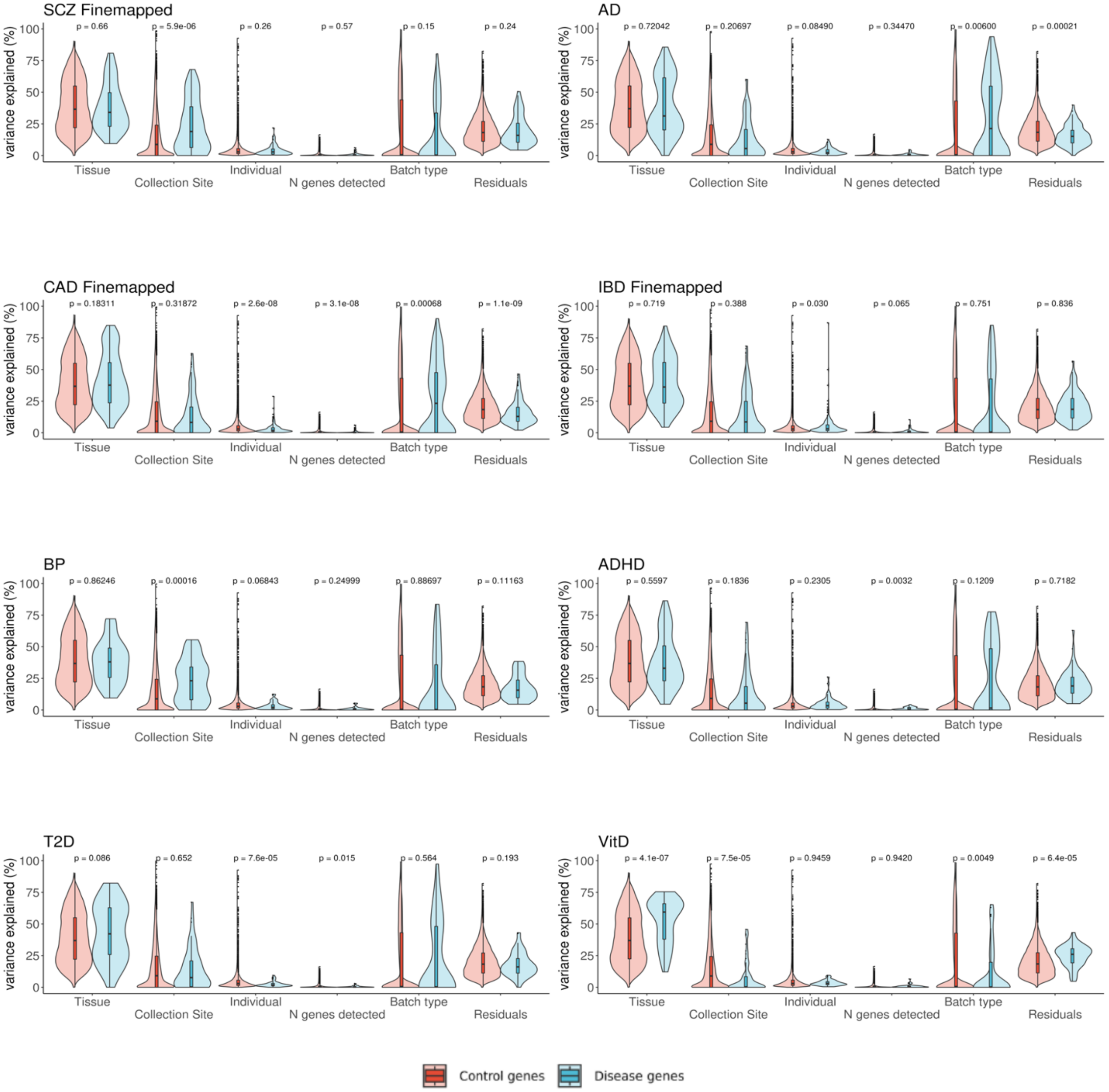


**Supplemental Figure 22.** Factors explaining gene expression in disease-associated genes via *Finemapping* vs other protein-coding genes. Every data point composing the violin plots represents the proportion of variance explained by a predictor (Tissue, Collection Site, Individual N genes detected, Batch Type) on the gene expression of an individual gene. This is calculated by fitting a linear mixed model for each gene. The results are color-coded to compare the variance explained by each factor in fine-mapped genes vs control genes.

### Biological and Technical Factors Influencing Disease Gene Expression (Variance Partition)

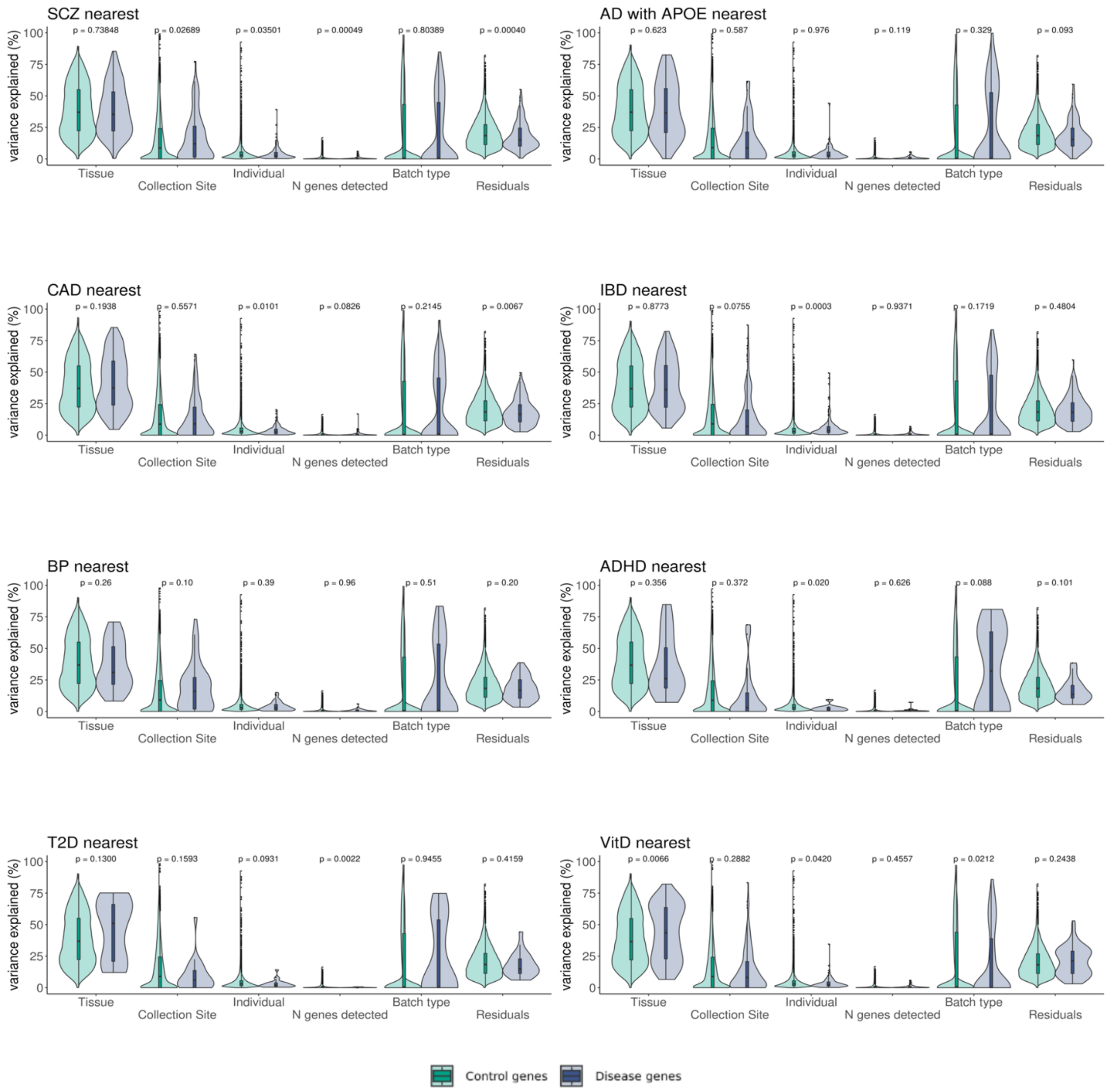


**Supplemental Figure 23.** Factors explaining gene expression in disease-associated genes via *nearest genomic location* vs other protein-coding genes. Every data point composing the violin plots represents the proportion of variance explained by a predictor (Tissue, Collection Site, Individual N genes detected, Batch Type) on the gene expression of an individual gene. This is calculated by fitting a linear mixed model for each gene. The results are color-coded to compare the variance explained by each factor in *nearest-to-GWAS-hit* genes vs control genes.

### Biological and Technical Factors Influencing Disease Gene Expression (Variance Partition)

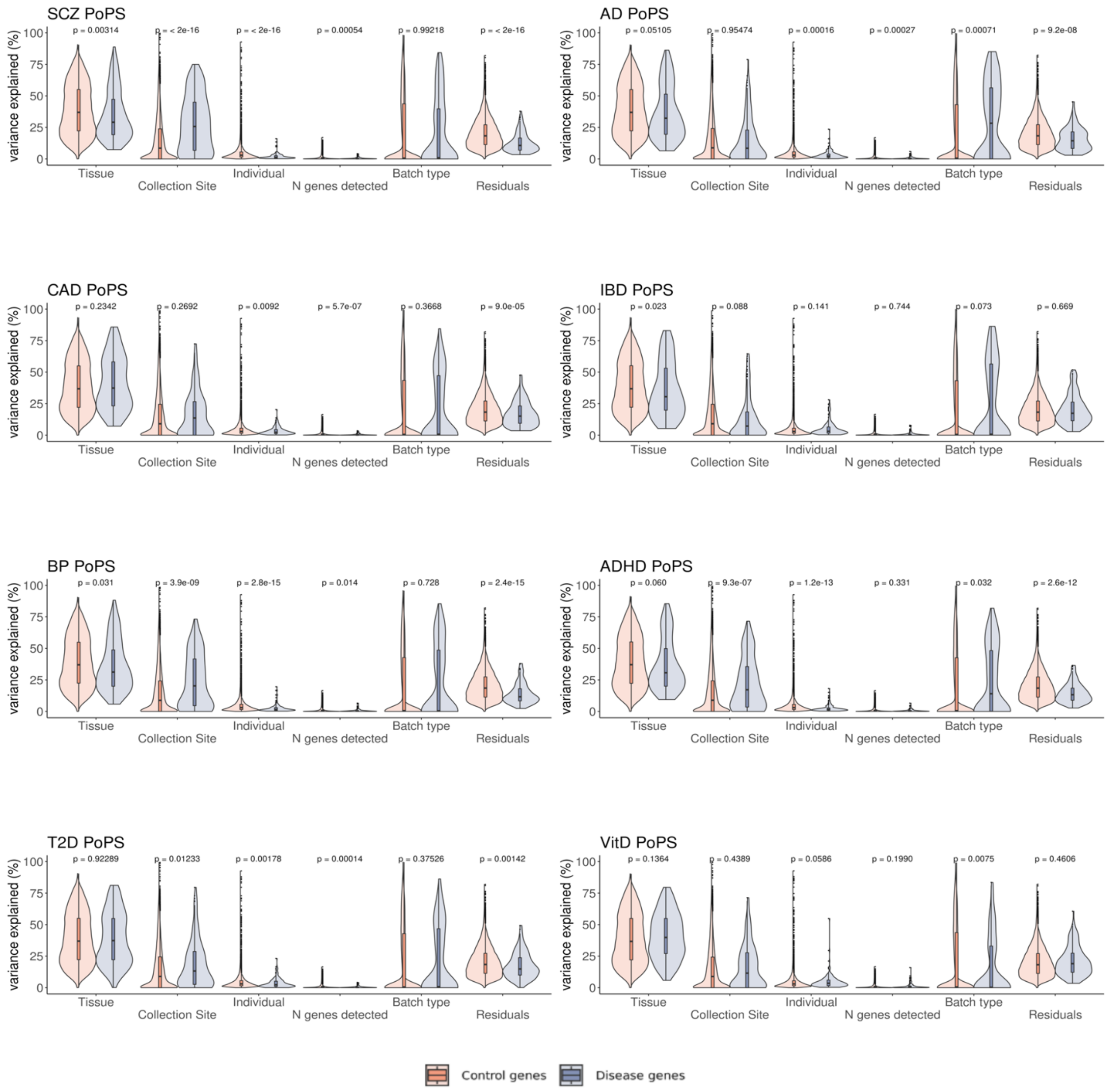


**Supplemental Figure 24.** Factors explaining gene expression in disease-associated genes via the PoPS prioritization method vs other protein-coding genes. Every data point composing the violin plots represents the proportion of variance explained by a predictor (Tissue, Collection Site, Individual N genes detected, Batch Type) on the gene expression of an individual gene. This is calculated by fitting a linear mixed model for each gene. The results are color-coded to compare the variance explained by each factor in *PoPS* genes vs control genes.
